# Assessing diagnostic accuracy of onchocerciasis rapid diagnostic tests using Bayesian latent class models

**DOI:** 10.64898/2026.01.20.26344416

**Authors:** Jayla Norman, N’Deye-Marie Bassabi-Alladjie, Pelagie M Boko-Collins, Dziedzom K. de Souza, Katherine Gass, Louise Hamill, Jerónimo Langa, Carson Moore, Nelvina Mucato, Rassul Nala, Sarah Sullivan, Emanuele Giorgi

## Abstract

**Background:** Onchocerciasis elimination programs increasingly rely on rapid diagnostic tests (RDTs) that detect antibodies to the Ov16 antigen, yet the performance of currently available rapid tests remains uncertain. The commercially available SD BIOLINE Ov16 RDT has shown inconsistent sensitivity when used on whole blood and does not consistently meet international performance thresholds for mapping or decisions to stop mass drug administration (MDA). Two novel onchocerciasis RDTs have been developed, DDTD RDT and GADx RDT, but their diagnostic accuracy has not been fully evaluated using methods that account for the absence of a true reference standard.

**Methodology/Principal Findings:** We pooled participant-level data from three field studies conducted in Mozambique, Ghana, and Benin in 2023 to evaluate two novel onchocerciasis RDTs and the commercially available SD BIOLINE Ov16 RDT. A Bayesian latent class model was used to estimate diagnostic sensitivity and specificity without assuming any test was a perfect reference standard. Across all model specifications, the GADx rapid test showed the highest sensitivity, with posterior median estimates (92.0%–92.8%) consistently above the 89% threshold recommended for decisions to stop mass drug administration (MDA). Posterior median sensitivity for the SD BIOLINE test was above this threshold in most models but below it under the field-based prior (83.2%). The DDTD RDT demonstrated lower sensitivity overall (86.6%–88.4%). Specificity estimates were consistently high for all tests but remained below the recommended threshold of 99.8% (highest median: 98.8%). Alternative definitions of a positive result for the multi-antigen DDTD RDT did not meaningfully alter its estimated performance.

**Conclusions/Significance:** In the study communities sampled, the two novel onchocerciasis RDTs generally yielded higher sensitivity estimates than the commercially available SD BIOLINE Ov16 RDT, with the GADx assay having the highest estimated sensitivity. However, posterior estimates for none of the evaluated tests reached the specificity threshold recommended to support decisions to stop MDA on their own. These findings highlight the need for improved diagnostic tools or complementary testing strategies to support elimination decision-making.

**Author Summary:** In this study, we evaluated three RDTs used to detect exposure to the parasite that causes river blindness. These tests are important tools for national programs that are working to eliminate the disease and determine when preventive treatment can be safely stopped. One challenge is that there is no perfect reference test against which new diagnostic tests can be evaluated. To address this, we pooled participant-level data from studies conducted in Mozambique, Ghana, and Benin and used a Bayesian latent class model to estimate test accuracy without assuming that any single test was perfect.

We found that one of the novel tests, the GADx RDT, identified people with exposure more accurately than the other evaluated tests, meeting the established WHO threshold for stopping MDA. The commercially available SD BIOLINE test and the DDTD test showed lower estimated accuracy than the GADx test. Although all three tests correctly identified most people who were not exposed, none achieved the very high level of specificity recommended to confidently support decisions to stop preventive treatment. As such, stop-MDA decisions should not rely on any of these RDTs alone. Further improvements, developments, and robust assessment are needed first.

Our findings suggest that while newer onchocerciasis RDTs are improving, additional advances are needed before these tests can independently support decisions to stop preventive treatment. These results provide evidence to inform the evaluation and selection of diagnostic tools as national programs progress from disease control toward onchocerciasis elimination.

## Introduction

Onchocerciasis, commonly known as river blindness, remains a major public health challenge in parts of Africa, Latin America, and Yemen. Caused by the filarial parasite *Onchocerca volvulus* and transmitted by the bite of infected blackflies (*Simulium* spp.), the disease is associated with severe morbidity, including debilitating skin disease, visual impairment, and blindness (1). Global control efforts have significantly reduced transmission in many endemic areas, but the World Health Organization (WHO) has shifted its focus from morbidity control toward elimination, setting ambitious targets for interruption of transmission by 2030. Achieving these goals requires robust tools for both mapping areas where the disease is still present and for making programmatic decisions about when to stop mass drug administration (MDA).

Current diagnostic methods for human onchocerciasis include the Ov16 rapid diagnostic test (RDT), which detects IgG4 antibodies against the Ov16 antigen, the diethylcarbamazine (DEC) patch test, skin snip microscopy, and enzyme-linked immunosorbent assay (ELISA)-based methods (2). Microscopic examination of skin snips has traditionally served as the reference standard for diagnosing and monitoring *O. volvulus* infection (3). However, the sensitivity of skin snip microscopy decreases as the density of microfilaria in the skin decreases (4). Detection of amplified parasite DNA in skin snips has been proposed as a solution for the low sensitivity of microscopy. Switching from microscopy to amplification of parasite DNA has generally been found to increase the sensitivity of skin snip assays and has become the standard for the diagnosis of patent *O. volvulus* infection in humans (3). Aside from being an insensitive indicator of infection, it is well documented that skin snips are painful and not well tolerated by the affected populations (5–8). Microscopy and DNA amplification both require specialized lab expertise, an additional limitation for many countries impacted by onchocerciasis. Lastly, as we shift from control to elimination, the insensitivity of skin snips poses a greater issue. A more sensitive indicator is needed because microfilaria can take years to develop in a person after they are exposed. Previous studies have shown that antibodies to Ov16 become detectable approximately 1-2 years before microfilariae appear in the skin (9).

Current onchocerciasis elimination strategies rely on assays that detect antibodies against the Ov16 antigen as indicators of exposure to *O. volvulus.* Renewed interest in Ov16 antibody detection has led to the development of two novel onchocerciasis RDTs intended to complement the commercially available SD BIOLINE Ov16 RDT (3). Interest in Ov16 RDTs reflects the need for a more sensitive indicator of exposure that is suitable for field use, supports elimination program decision-making, and does not require specialized laboratory expertise or equipment. While Ov16 RDTs are appealing because of their field applicability and ease of use, the WHO, through its Diagnostic Technical Advisory Group, has published a Target Product Profile (TPP) for the diagnostic needs for onchocerciasis, which explicitly calls for diagnostics with at least 60% sensitivity for mapping purposes, at least 89% sensitivity for decisions to stop MDA, and at least 99.8% specificity regardless of intended use (10). These benchmarks are particularly important in elimination settings, where false-negative results may allow ongoing transmission to go undetected, whereas false-positive results may lead to unnecessary continuation of MDA.

Currently, the SD BIOLINE Ov16 RDT is the only commercially available Ov16 antibody RDT and it is produced by Standard Diagnostic (SD). However, the SD BIOLINE Ov16 RDT has demonstrated lower performance on fresh whole blood as it does on blood eluted from dried blood spots (DBS) (11–14). Consequently, onchocerciasis elimination programs are currently collecting DBS in the field and then eluting them in a lab to run on the RDTs (14). This creates barriers including the addition of complexity, time, and cost to the surveys. Furthermore, the SD BIOLINE Ov16 RDT does not meet the standards outlined in the TPP, even when run from DBS. Its sensitivity and specificity are approximately 81% and 99%, respectively (14–16). Despite a recent scale-up in the use of Ov16 RDTs, questions remain about whether the currently available RDT meets these thresholds across different epidemiological contexts. Reported sensitivity and specificity estimates vary by study population, geographic setting, and reference standard, and few evaluations have incorporated modeling approaches to account for the imperfect nature of “gold standards” in parasitological diagnosis. Bayesian latent class models have been used to address this problem for other parasitic and infectious diseases lacking a reference standard, including schistosomiasis, Chagas disease, tuberculosis, and canine visceral leishmaniasis (27–29), but this approach has not yet been applied to Ov16 based diagnostics for onchocerciasis. In elimination settings where microfilaria prevalence is very low, traditional parasitological methods (e.g., skin snip microscopy) lack sensitivity, further complicating the assessment of RDT accuracy (3, 17). Furthermore, the evaluated diagnostic methods target different biological indicators of infection or exposure. Skin snip-based methods detect the presence of parasites in the skin, whereas Ov16 RDTs detect host antibody responses following exposure to *O. volvulus* larvae (18). For these reasons, an investment in developing new RDTs has emerged. Notably, two novel onchocerciasis RDTs by Drug & Diagnostics for Tropical Diseases (DDTD) and Global Access Diagnostics (GADx) have been developed. The GADx RDT is an Ov16 RDT, while the DDTD RDT is a multi-antigen RDT which detected antibodies against the Ov16 antigen (test line 1, T1) and OvOC3261 (test line 2, T2).

The objective of this study was to estimate the diagnostic sensitivity and specificity of two novel onchocerciasis RDTs (DDTD and GADx) and the commercially available SD BIOLINE Ov16 RDT using pooled participant-level data from three coordinated field studies conducted in Mozambique, Ghana, and Benin. Because no single diagnostic method can be considered a perfect reference standard for measuring exposure to *O. volvulus,* we applied Bayesian latent class models to estimate test performance without assuming that any evaluated test was perfect. We further compared the estimated diagnostic performance of each RDT with WHO Target Product Profile thresholds for mapping and decisions to stop MDA. These findings provide evidence to inform programmatic decision-making as endemic countries progress towards the elimination of onchocerciasis.

To our knowledge this is the first evaluation of these two novel onchocerciasis rapid diagnostic tests alongside the commercially available test across multiple countries using a Bayesian latent class framework, which estimates diagnostic accuracy, including specificity, without relying on any single test as a reference standard.

## Methods

### Data

This study pooled harmonized participant-level data from three coordinated multi-country field evaluations of onchocerciasis RDTs conducted in Mozambique, Benin, and Ghana in 2023; the maps of the study sites from each country are shown in Figure 1S in S1 Appendix. Each evaluation assessed the performance and feasibility of multiple onchocerciasis RDTs among individuals aged ≥5 years in historically onchocerciasis-endemic communities. Data from each country evaluation were combined into a single analytical dataset to estimate average diagnostic sensitivity and specificity across the included field settings. Details of study implementation, including community selection and data collection timelines, are described in the subsequent section below. Additional details about sampling methods and procedures by country are available in Table S2 in S1 Appendix.

**Fig 1.**
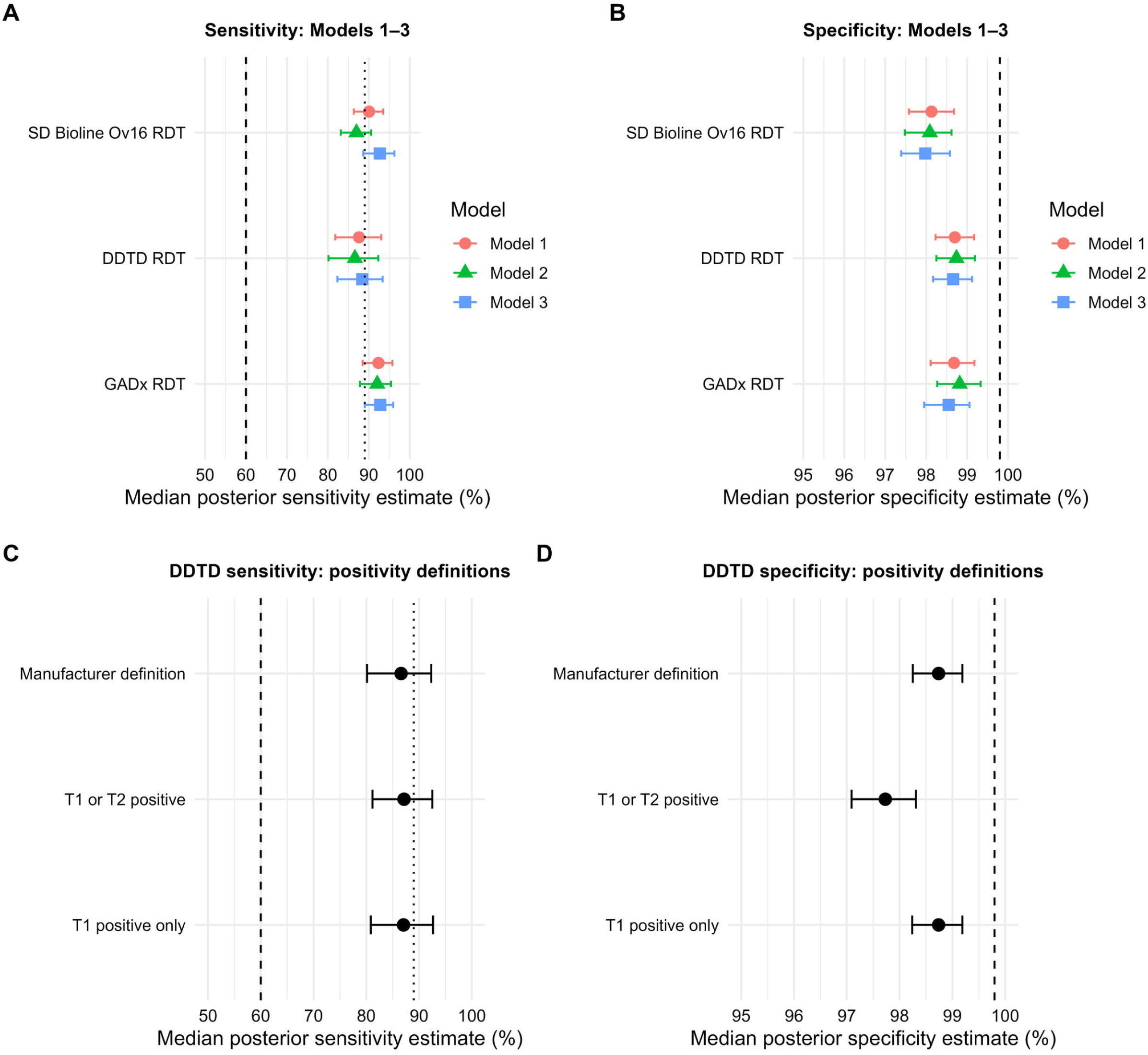
Posterior estimates of sensitivity and specificity across Bayesian latent class models. Points represent posterior median estimates, and horizontal lines represent 95% credible intervals (CrIs). Panels A and B show posterior median estimates of sensitivity and specificity, respectively, for the SD BIOLINE Ov16, DDTD, and GADx RDTs under Models 1-3. Panels C and D show posterior median estimates and 95% CrIs for DDTD sensitivity and specificity, respectively, under the manufacturer-defined positivity criterion (Model 2) and alternative positivity definitions evaluated in Models 4 and 5. Vertical dashed lines indicate the WHO target product profile thresholds of ≥60% sensitivity for mapping and ≥99.8% specificity, while vertical dotted lines in the sensitivity panels indicate the ≥89% sensitivity threshold for stopping MDA. In panels A and B, colors and shapes distinguish Models 1-3, as indicated in the figure legend; panels C and D display the DDTD positivity definitions directly on the y-axis.

Raw datasets from each country were reviewed for consistency and completeness. Variable names and coding were standardized, and duplicate or incomplete records were removed before analysis. SD Bioline had no invalid results at any site. DDTD had 14 invalid results overall (3 in Mozambique, 3 in Benin, 8 in Ghana) and GADx had 5 (2 in Mozambique, 2 in Benin, 1 in Ghana); reasons included blood discoloration obscuring the results window and device-related strip damage in Mozambique, and absent control lines in Benin. Reasons for Ghana’s invalid results were not recorded. Invalid and missing results were treated as missing at the individual-test level; participants with a result for at least one test contributed to the model, and only participants with no valid result on any test were excluded from analysis. Data from all sites were harmonized for combined analysis. All data management and analysis were conducted in R version 4.4.1 (17).

### Study Populations and Sampling Procedures

#### Mozambique

Data were collected between September 18-25, 2023, in two communities: Namarrenda (Milange District) and Nantuto (Molumbo District). The study team conducted sensitization meetings with district health officials in both study districts before data collection. All residents aged 5 years and older were eligible to participate. Communities were selected based on accessibility within the ongoing DISSECT survey (19). A total of 409 individuals were enrolled.

#### Benin

Fieldwork occurred between July 23-August 5, 2023, in three districts: Tchaourou, Savè, and Bassila. Community sensitization sessions were conducted with leaders in the selected districts before field activities. Sampling initially targeted two districts, but Bassila was added when enrollment targets could not be met in Tchaourou. Households were selected based on the presence of at least one child aged 5-9 years, as they were the target population of Benin’s CDC-funded study. A total of 948 individuals were enrolled.

#### Ghana

Data were collected between September 24-October 20, 2023, across seven communities in three districts (Adaklu, Hohoe, Nkwanta North). Community sensitization sessions were conducted before field activities. All residents aged ≥5 years were eligible. A total of 1,700 individuals enrolled.

### Ethics Statement

All studies contributing data to this analysis obtained ethical approval from their respective institutional review boards. Written informed consent was obtained from all adult participants, and parental consent with child assent was obtained for participants under 18 years of age.

### Statistical Analysis

Latent class models (LCMs) have been widely used for evaluating diagnostic test accuracy in the absence of a gold standard test (20, 27–29). These approaches typically rely on the assumption of *conditional independence*, whereby the results of multiple diagnostic tests are assumed to be independent of one another after conditioning on the true, but unobserved, disease status (20–22). Within this framework, a Bayesian LCM was used to jointly estimate the sensitivity and specificity of each Ov16 RDT, while accounting for the absence of a gold standard. This approach treats the true infection status of participants as an unobserved (latent) variable and infers test characteristics by integrating information from all available tests. The Bayesian latent class model also accommodated partial missingness at the individual-test level, so participants with a valid result for one or two tests contributed the information they had.

More details on the LCM are provided in S2 Appendix.

### Index Tests and Reference Methods

Three Ov16 RDTs were evaluated: SD BIOLINEOv16 RDT manufactured by Abbott, DDTD multi-antigen onchocerciasis antibody RDT, hereafter referred to as DDTD RDT, manufactured by Drug & Diagnostics for Tropical Diseases, and GADx Ov16 RDT, hereafter referred to as GAD, manufactured by Global Access Diagnostics. The two novel RDTs, DDTD RDT and GADx RDT, were performed on finger-prick whole blood samples following the manufacturer’s instructions for use, and the existing SD BIOLINEOv16 RDT was tested on eluted DBS. The DDTD RDT has two test lines (T1 and T2) that detect antibodies against different *O. volvulus* recombinant antigens (23, 24). The T1 test line of the DDTD RDT is used to indicate the presence of the Ov16 antigen, and the T2 test line is used to indicate the presence of the OVOC3261 antigen. According to the manufacturer’s instructions for use, a positive DDTD RDT result requires both the T1 and T2 test line to be positive. Alternative positivity definitions (T1 or T2 positive; T1 positive only) were assessed separately in sensitivity analyses (Models 4 and 5), as described below in the model specifications below.

To standardize the performance of the DDTD RDT and GADx RDT, as well as compare them to the currently recommended diagnostic approach, the SD Bioline Ov16 RDT served as the comparator of primary programmatic interest. However, because no test can be treated as a gold standard for Ov16 serology, all three tests were modelled symmetrically within a Bayesian latent class framework, with no test assumed perfect.

To assess the robustness of our findings, we considered five model specifications. An informative prior on at least one test (here, SD Bioline) helps ensure identifiability of the two latent classes and reduces the risk of label-switching during MCMC sampling (20). Hence, we assigned informative priors to SD Bioline, an established assay with independently reported performance data, while retaining noninformative priors for the two novel RDTs. Because manufacturer estimates are typically generated under near-ideal laboratory conditions and may not reflect performance in the field, we considered both manufacturer-reported and field-derived priors for SD Bioline, alongside a model with no prior information, to examine how sensitive our conclusions were to this choice. We additionally evaluated whether alternative definitions of a positive DDTD result affected its estimated accuracy.

In summary, we consider five model specifications, each considering different prior specifications or alternative definitions of a positive result.:

1. **Model 1:** Prior information on sensitivity and specificity of SD BIOLINE Ov16 RDT based on Abbott’s reporting of test performance (15, 16).
2. **Model 2:** Prior information on sensitivity and specificity of SD BIOLINE Ov16 RDT based on field estimates from previous research (5).
3. **Model 3:** No prior information specified on SD BIOLINE Ov16 RDT performance.
4. **Model 4:** Same prior information as in Model 2. Considers an alternative definition of positive test result for DDTD RDT where positive on either test line (T1 or T2) is defined as positive. For reference, manufacturer instructions require both test lines to be positive to conclude a positive test result.
5. **Model 5:** Same prior information as in Model 2. Considers an alternative definition of positive test result for DDTD RDT where positive on T1 test line only is defined as positive. For reference, manufacturer instructions require both test lines to be positive to conclude a positive test result.

Models 1-3 were the main models of analysis for evaluating the performance of the two novel RDTs. Models 4 and 5 were complementary models that were developed to evaluate if alternative definitions of a positive test for the DDTD RDT impacted its performance.

Posterior distributions of sensitivity and specificity were estimated using Markov Chain Monte Carlo (MCMC) simulation with 10,000 iterations, with the first 4,000 discarded as burn-in. The remaining 6,000 iterations were used to generate posterior distributions of test sensitivity, specificity, and infection prevalence. Aside from Model 3, noninformative uniform priors on the interval (0,1) were specified for all parameters to minimize assumptions about test performance. Posterior convergence was verified using trace plots and the Gelman-Rubin statistic, with R < 1.1 considered acceptable.

Sensitivity estimates were compared against the WHO thresholds for mapping (≥60%) and for stopping MDA (≥89%), and specificity estimates were compared against the WHO threshold of ≥99.8%. Results are presented as posterior medians with 95% credible intervals.

## Results

A total of 3,057 persons were enrolled across all sites, with 3,040 persons contributing to the pooled analysis, and 2,743 complete observations. These 3,040 individuals represent persons who had data for at least one of the three tests. Missing or invalid test results were incorporated into the model as missing.

### Posterior Sensitivity and Specificity Estimates

Across all five Bayesian latent class models, posterior estimates of sensitivity and specificity varied by test. Table 1 presents posterior median estimates and 95% credible intervals (CrIs) for prevalence, sensitivity, and specificity under Models 1-3. Posterior sensitivity and specificity estimates for the three RDTs across these models are also presented graphically in Fig 1A and 1B.

**Table 1.**
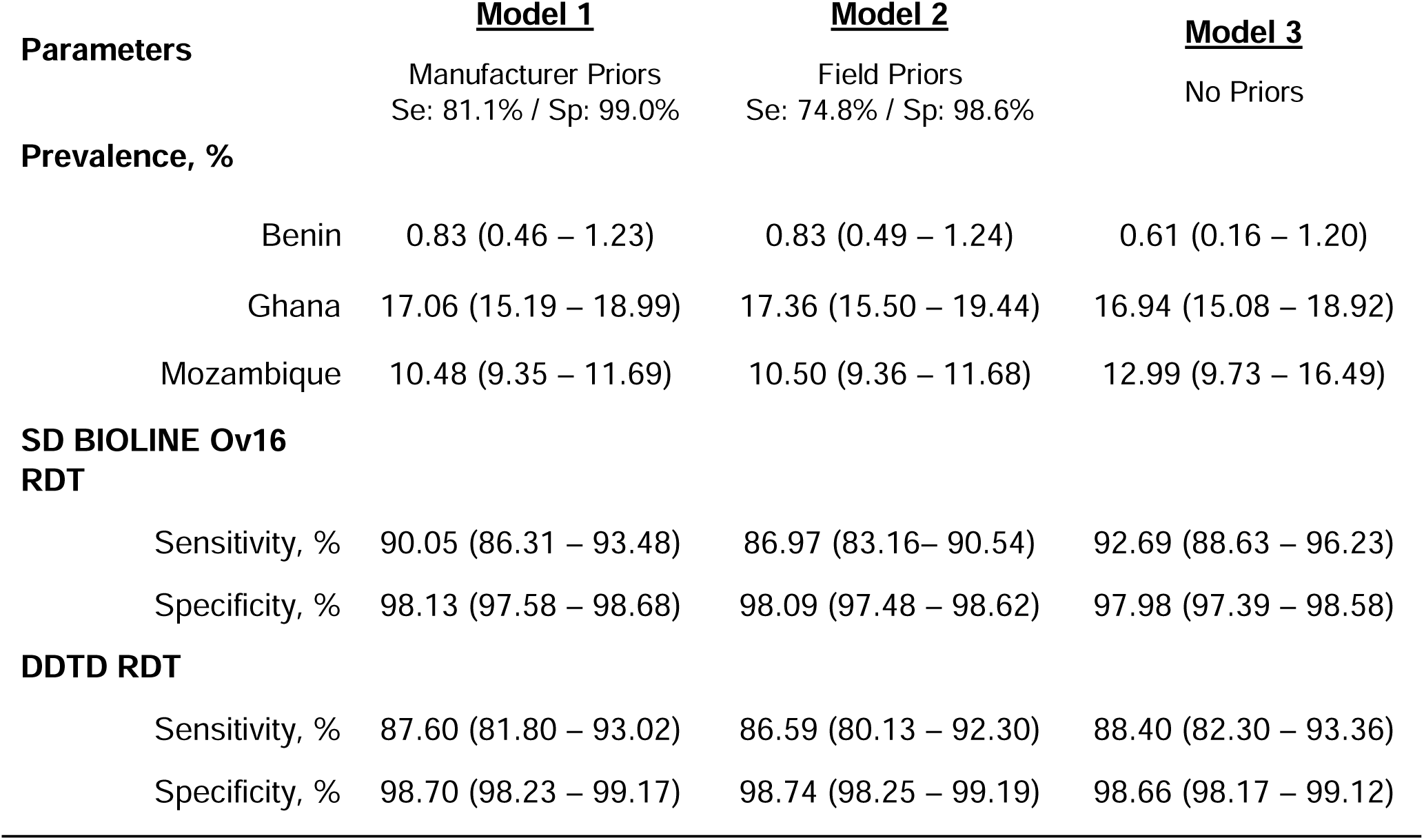
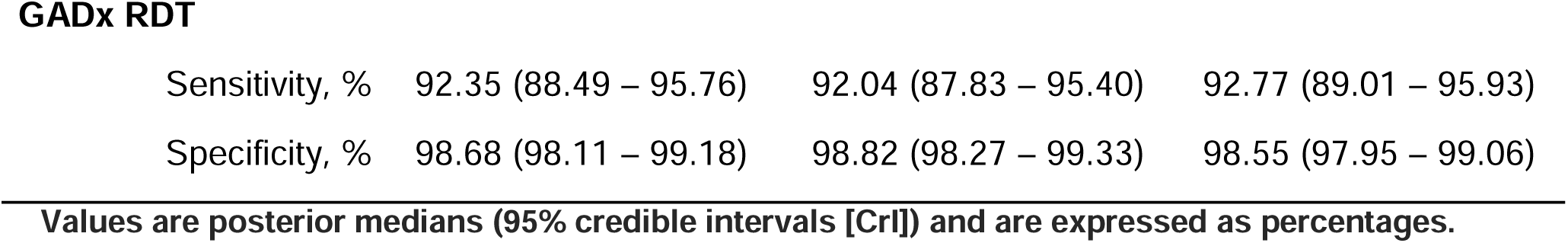
Posterior estimates of prevalence and diagnostic performance under Models 1-3.

The GADx RDT demonstrated the highest sensitivity, exceeding WHO’s 89% stopping threshold in all three models (Model 1: 92.4%, 95% CrI: 88.5 - 95.8; Model 2: 92.0%, 95% CrI: 87.8 – 95.4; Model 3: 92.8%, 95% CrI: 89.0 – 95.9).

The SD BIOLINE Ov16 RDT met the 89% threshold in Models 1 and 3 but fell slightly below in Model 2 (Model 1: 90.1%, 95% CrI: 86.3 – 93.5; Model 2: 87.0%, 95% CrI: 83.2 – 90.5); Model 3: 92.7%, 95% CrI: 88.6 – 96.2).

The DDTD RDT demonstrated lower sensitivity across all models (Model 1: 87.6%, 95% CrI: 81.8 – 93.0; Model 2: 86.6%, 95% CrI: 80.1 – 92.3; Model 3: 88.4%, 95% CrI: 82.3 – 93.4), with point estimates averaging approximately 1.5 percentage points below the 89% threshold.

Specificity estimates were uniformly high across all tests but fell short of WHO’s ≥ 99.8% threshold. The highest posterior median specificity was observed for GADx (Model 2: 98.8%, 95% CrI: 98.3 – 99.3), followed closely by DDTD (98.7%, 95% CrI: 98.3 – 99.2) and SD BIOLINE (98.1%; 95% CrI: 97.5 – 98.6).

Table 2 summarizes posterior estimates under the alternative definitions of positivity of the DDTD RDT evaluated in Models 2, 4, and 5; corresponding DDTD sensitivity and specificity estimates are shown graphically in Fig 1C and 1D. The alternative positivity definitions resulted in relatively stable sensitivity estimates across the three models, whereas specificity was lower when positivity was defined as either test line positive (Model 4). Median posterior specificity estimates for the DDTD RDT in Models 2 and 5 were equivalent (98.74%) with almost identical 95% CrIs, while Model 4’s median estimate was slightly lower at 97.73%; sensitivity was approximately 87% in the three models.

**Table 2.**
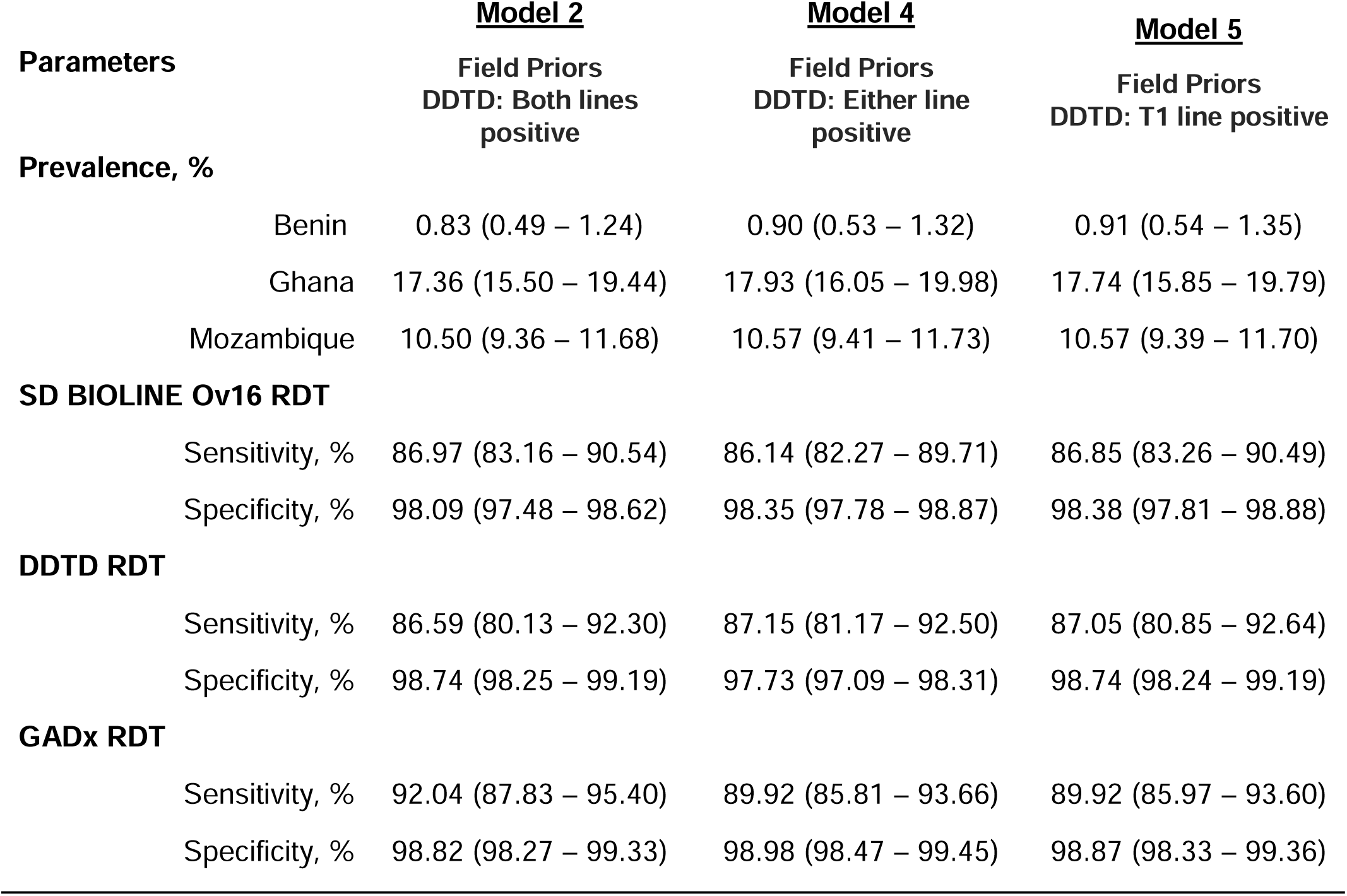
Posterior estimates under alternative DDTD positivity definitions (Models 2, 4, and 5).

### Values are posterior medians (95% credible intervals [CrI]) and are expressed as percentages

To provide decision-relevant summaries beyond posterior medians, we calculated the posterior probability that each test met the WHO thresholds for sensitivity and specificity, together with expected false positives per 1,000 tested and predictive values across a range of prevalences (Tables A1a–e). The probability that the GADx RDT met the 89% sensitivity threshold was consistently high across models (67–97%), reflecting stable estimates under alternative priors and the DDTD RDT positivity definitions, whereas the corresponding probability for SD Bioline Ov16 RDT varied substantially (5–96%) and tracked directly with the choice of informative prior in Models 1–3. DDTD’s probability of meeting the sensitivity threshold remained consistently low (21–42%) regardless of specification. No test had an appreciable posterior probability of meeting the 99.8% specificity threshold in any model. Expected false positives per 1,000 tested at 1% prevalence were broadly stable for SD Bioline (16–20) and GADx (10–14) across models, but rose for DDTD under the alternate either-line positivity definition (Model 4: 22.5, versus 12.5–13.3 under the manufacturer definition), consistent with the corresponding drop in DDTD specificity in that model. These patterns indicate that heterogeneity in decision-relevant metrics across models was driven primarily by the choice of prior for SD Bioline and by the DDTD positivity definition, rather than by underlying differences in novel-test performance.

## Discussion

This study evaluated the diagnostic performance of three onchocerciasis RDTs using Bayesian latent class models, providing estimates of sensitivity and specificity in the absence of an accurate reference standard. Across all models, the GADx RDT consistently met WHO’s 89% sensitivity threshold for decisions to stop MDA, while SD BIOLINE Ov16 RDT met this threshold in most but not all model specifications. In contrast, the DDTD RDT fell slightly below the threshold across all models. Importantly, none of the evaluated RDTs achieved the WHO specificity benchmark of ≥ 99.8% (*see Table 1)*, indicating that they may not yet be suitable for standalone use in decisions to stop MDA.

Our findings align with prior evaluations of Ov16 RDTs, which have reported variable sensitivity and specificity across field and laboratory settings. However, to the best of our knowledge, none of the related evaluations of these RDTs have employed latent class models to account for imperfect reference standards. By incorporating Bayesian LCMs, our analysis provides more robust estimates and avoids the bias that arises when assuming any single test is a gold standard. Two recent studies have evaluated overlapping sets of these RDTs using different approaches. A site level analysis of the Ghana field data compared the DDTD and GADx tests against SD Bioline as a reference standard, within a non-inferiority framework (25). Because skin snip qPCR identified too few positives in that setting, specificity could not be estimated for either novel test. A separate evaluation conducted in pregnant and postpartum women in South Sudan compared all three RDTs against several reference standards, including skin snip microscopy, O150 qPCR, and Ov16 ELISA (26). Both approaches rest on the assumption that at least one comparator, whether SD Bioline or a parasitological or molecular reference test, correctly classifies true exposure status, an assumption that is difficult to justify given the well documented limitations of each of these tests in low transmission settings (3, 4, 14, 26). The present analysis overcomes this reliance entirely. By treating all three RDTs symmetrically within a Bayesian latent class framework and pooling data across three countries, we estimate sensitivity and specificity for every test without designating any single test or reference method as the standard against which the others are judged. This analytical approach has allowed us to show that none of the three tests currently meets the WHO threshold for stopping MDA decisions, a finding that the reference standard dependent designs of the previous studies were not positioned to establish.

These findings have important implications for onchocerciasis elimination programs. While the GADx RDT’s sensitivity suggests it could support decisions to stop MDA in some settings, the shortfall in specificity for all tests raises concerns about false-positive results. Reliance on tests with imperfect specificity could lead to the error of falsely continuing to treat areas that need it. This shortfall also impacts the ability to map onchocerciasis. As outlined by the WHO (10), a sensitive and specific test is required to map onchocerciasis in hypo-endemic areas and overcome the limitations of existing tools. While these novel RDTs did fare better than current diagnostic tools, representing improvement and innovation in the field, they do not completely solve existing diagnostic challenges. Highly specific diagnostic tests are essential for accurately determining the need for MDA and ensuring that elimination efforts are effective and sustainable. These results highlight the need for either improved test specificity or confirmatory testing algorithms before these RDTs can reliably support stopping decisions, mapping, and other programmatic needs.

### Limitations

We discuss in turn several important limitations of this study, arising from both data constraints and limitations of existing methodology.

All models assumed conditional independence between tests given latent exposure status. The three assays differ in antigen target, platform, and specimen pathway. The two novel RDTs were performed on whole blood at the point of care, while the SD Bioline RDT was performed on dried blood spots eluted separately in the laboratory. Shared operational sources of correlated error are therefore limited. A residual source of dependence nonetheless cannot be excluded, since all three tests ultimately detect the same underlying host IgG4 antibody response and individuals with titres near the detection threshold could plausibly fail more than one test together. Any such dependence is expected to be positive, and positive conditional dependence causes latent class models to overstate both sensitivity and specificity (20). Our estimates should therefore be read as upper bounds on true performance, which means our central finding, that no evaluated test reaches the WHO specificity threshold, is conservative rather than at risk from this assumption. We did not attempt a dependent latent class model in this context. With only three tests per participant, the model is already weakly identified, which is why an informative prior on one test was used to help stabilize estimation. Adding dependence parameters would likely exacerbate this identifiability problem, which is one of the main reasons why addressing violations of conditional independence remains an open methodological issue for latent class models in this context and warrants further research.

The specification of priors for the SD Bioline test also influenced its estimated performance. We compared three prior specifications for this test, drawing in turn on manufacturer reported performance, on field derived estimates from previous research, and on no external information at all. Sensitivity for SD Bioline was lowest under the field derived prior, whose prior mean sits well below the values reported by the manufacturer, and highest when no external information was imposed. The two novel tests, which carried noninformative priors throughout, produced essentially identical estimates under all three specifications. The variability observed for SD Bioline therefore reflects differences in which body of external evidence is treated as most credible for that test rather than instability of the analytical approach and comparing the three specifications side by side is itself the robustness check we performed. It also means results from SD Bioline should be read with more caution than those about the two novel tests.

Test accuracy was assumed common across the three countries while prevalence was allowed to vary by site. It is reasonable to assume that sensitivity and specificity are properties of the assay, defined conditionally on true exposure status, and that identical test lots and protocols were used throughout, so we regard this as a defensible starting assumption. If this assumption is violated, and true performance differs across these epidemiological and operational settings, the pooled estimates would obscure that heterogeneity, in a direction that cannot be predicted a priori. Additionally, country specific accuracy estimates raise further issues. Such estimates would require enough latently exposed participants within each site to inform sensitivity, and this requirement is not met in every setting. In Benin, estimated prevalence is below 1%, so the number of exposed individuals in that site is too small to support a local sensitivity estimate; such an estimate would be determined largely by its prior rather than by the observed data. Estimation of specificity, instead, would not suffer from this issue, since unexposed participants are numerous at every site. Hierarchical extensions that partially pool accuracy across sites remain an important direction for future methodological work.

Differences in specimen type represent a further limitation. Because each assay was implemented under its own intended workflow, with the novel tests run on finger prick whole blood in the field and SD Bioline run on dried blood spots eluted in the laboratory, specimen type is confounded with assay. Differences in performance between SD Bioline and the novel tests cannot be attributed to test chemistry alone, and the present data do not allow specimen related effects to be separated from assay related ones. Our estimates are best interpreted as describing performance as each test would be deployed in practice.

Finally, the contributing data were generated by operational research studies designed to assess field acceptability and non-inferiority of the novel tests relative to SD Bioline as a working comparator, rather than to estimate diagnostic accuracy in the absence of a reference standard, which is the objective of the present analysis. The ability of the novel tests to utilize whole blood in field settings, rather than diagnostic accuracy per se, was the primary outcome these studies were designed to demonstrate, and sample sizes and data collection procedures were accordingly optimized for operational feasibility rather than for precision in sensitivity and specificity estimation. A related constraint is that the latent class framework treats each test as a single binary result, so it cannot evaluate whether the multiple antigen targets of the DDTD test carry incremental diagnostic information beyond the overall positive or negative classification. We examined alternative definitions of positivity for this test and found its estimated performance largely unchanged, but a design that exploits the antigen specific lines directly would require a different modelling approach.

### Future Directions and Conclusion

Future work should prioritize improving the specificity of Ov16-based diagnostics or exploring new antigens that maintain high sensitivity while reducing false positives. Additional studies using longitudinal data and larger sample sizes could refine performance estimates and assess how test characteristics vary across epidemiological settings. Incorporating confirmatory algorithms or multi-test strategies may also offer a path forward for programs that are nearing elimination targets but require robust evidence before halting interventions.

In summary, this analysis provides critical evidence about the current capabilities and limitations of Ov16 RDTs in onchocerciasis elimination programs. While certain RDTs demonstrate sufficient sensitivity to inform some programmatic decisions, the shortfall in specificity remains a key barrier to their use as stand-alone tools for stopping MDA. Continued innovation and careful validation of diagnostic tools will be essential for achieving WHO’s 2030 elimination goals.

## Supporting information

S_trace_plots (referenced in Supplemental Appendix 1)

## Data Availability

Data for each country are available at the following Dataverse DOIs: Benin - https://doi.org/10.15139/S3/RFZ4BE, Ghana - https://doi.org/10.15139/S3/MP1EJX, Mozambique - https://doi.org/10.15139/S3/ZOXQHS.

