## Supplemental Appendix 1 for "Assessing diagnostic accuracy of onchocerciasis rapid diagnostic tests using Bayesian latent class models"

**S1 Appendix. Additional figures and tables**

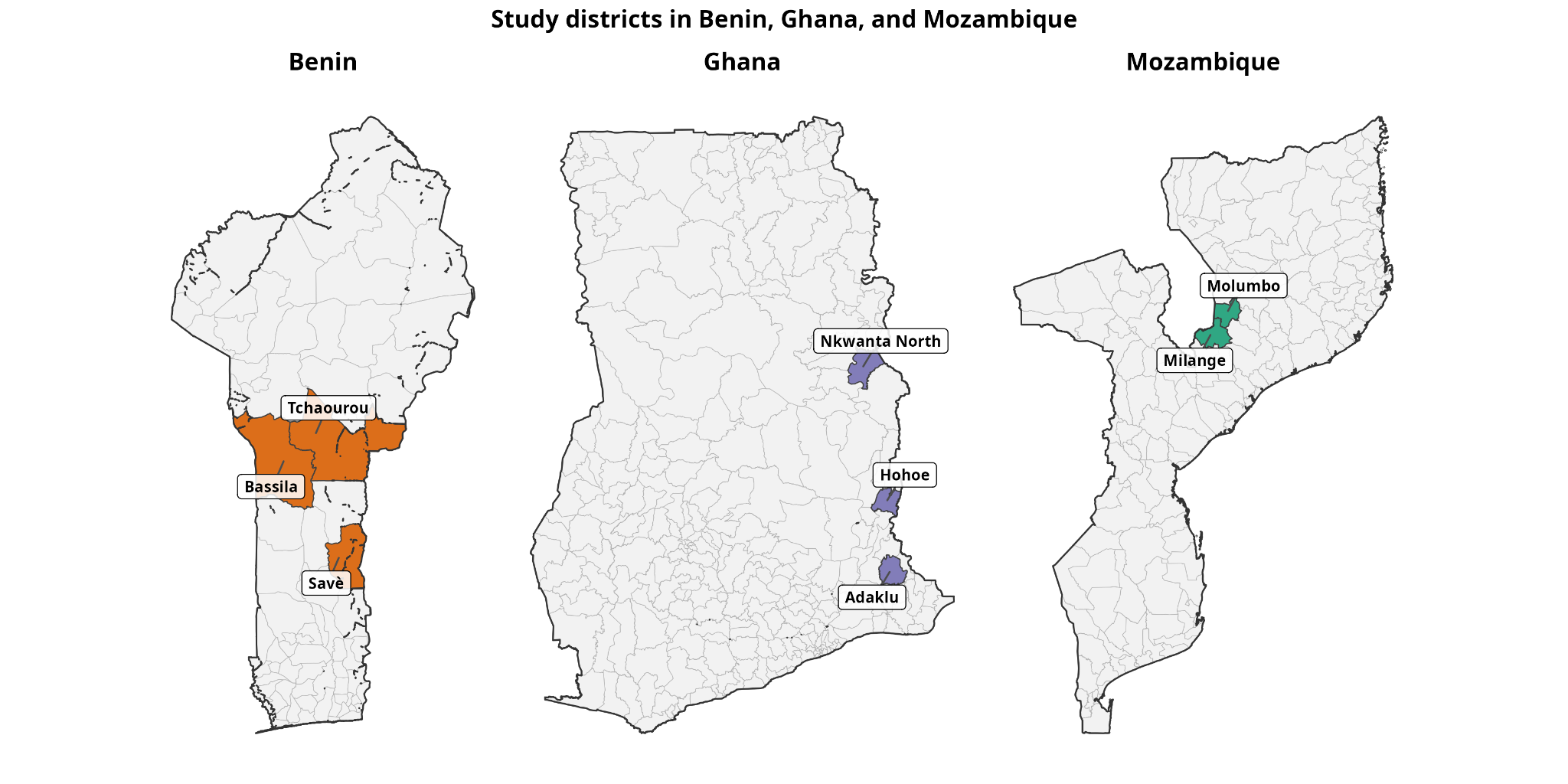

**Fig 1S. Study districts included in the pooled analysis.** District (admin level 2) boundaries for Benin (left), Ghana (center), and Mozambique (right), with the districts contributing participant-level data to this analysis shaded and labelled. Benin: Tchaourou, Savè, and Bassila districts. Ghana: Adaklu, Hohoe, and Nkwanta North districts. Mozambique: Milange and Molumbo districts (Zambézia Province). Unshaded districts are shown for national context only and did not contribute data to this study. Country boundaries and district boundaries for Benin and Ghana are from the Database of Global Administrative Areas (GADM), version 4.1; district boundaries for Mozambique are from geoBoundaries (Open Database License), reflecting the current administrative structure including the post-2013 division of Milange and Molumbo districts.

**Table S1a. Model 1 – Decision metrics for Ov16 RDTs**

Posterior median sensitivity (Se) and specificity (Sp), probability of meeting WHO thresholds (Se ≥89% for stop-MDA decisions, Sp ≥99.8%), predictive values (PPV, NPV) at representative elimination-setting prevalences, and expected number of false positives per 1,000 tested.

| Test | Median Se (%) | P(Se ≥89%) | Median Sp (%) | P(Sp ≥99.8%) |
| --- | --- | --- | --- | --- |
| SDBioline | 90.1 | 0.709 | 98.1 | 0.000 |
| DDTD | 87.6 | 0.315 | 98.7 | 0.000 |
| GADx | 92.4 | 0.946 | 98.7 | 0.000 |
| Prevalence | **Test** | **PPV (%)** | **NPV (%)** | |
| 0.6% | DDTD | 28.8 | 99.9 | |
| 0.6% | GADx | 29.6 | 100.0 | |
| 0.6% | SDBioline | 22.5 | 99.9 | |
| 1.1% | DDTD | 42.7 | 99.9 | |
| 1.1% | GADx | 43.6 | 99.9 | |
| 1.1% | SDBioline | 34.8 | 99.9 | |
| 2.1% | DDTD | 59.0 | 99.7 | |
| 2.1% | GADx | 59.9 | 99.8 | |
| 2.1% | SDBioline | 50.7 | 99.8 | |
| 5.1% | DDTD | 78.3 | 99.3 | |
| 5.1% | GADx | 78.9 | 99.6 | |
| 5.1% | SDBioline | 72.1 | 99.5 | |
| 10.1% | DDTD | 88.3 | 98.6 | |
| 10.1% | GADx | 88.7 | 99.1 | |
| 10.1% | SDBioline | 84.4 | 98.9 | |
| Prevalence | **Test** | **Expected FP per 1000** | **95% CrI** | |
| 1% | SDBioline | 18.5 | 13.5 – 24.5 | |
| 5% | SDBioline | 17.8 | 12.9 – 23.5 | |
| 10% | SDBioline | 16.9 | 12.3 – 22.3 | |
| 1% | DDTD | 12.9 | 8.7 – 18.1 | |
| 5% | DDTD | 12.4 | 8.4 – 17.3 | |
| 10% | DDTD | 11.7 | 7.9 – 16.4 | |
| 1% | GADx | 13.1 | 8.2 – 18.9 | |
| 5% | GADx | 12.6 | 7.9 – 18.1 | |
| 10% | GADx | 11.9 | 7.5 – 17.1 | |

**Table S1b. Model 2 – Decision metrics for Ov16 RDTs**

Posterior median sensitivity (Se) and specificity (Sp), probability of meeting WHO thresholds (Se ≥89% for stop-MDA decisions, Sp ≥99.8%), predictive values (PPV, NPV) at representative elimination-setting prevalences, and expected number of false positives per 1,000 tested.

| Test | Median Se (%) | P(Se ≥89%) | Median Sp (%) | P(Sp ≥99.8%) |
| --- | --- | --- | --- | --- |
| SDBioline | 87.0 | 0.136 | 98.1 | 0.000 |
| DDTD | 86.6 | 0.208 | 98.7 | 0.000 |
| GADx | 92.0 | 0.924 | 98.8 | 0.000 |
| Prevalence | **Test** | **PPV (%)** | **NPV (%)** | |
| 0.6% | DDTD | 29.3 | 99.9 | |
| 0.6% | GADx | 32.0 | 100.0 | |
| 0.6% | SDBioline | 21.5 | 99.9 | |
| 1.1% | DDTD | 43.3 | 99.8 | |
| 1.1% | GADx | 46.4 | 99.9 | |
| 1.1% | SDBioline | 33.5 | 99.9 | |
| 2.1% | DDTD | 59.5 | 99.7 | |
| 2.1% | GADx | 62.5 | 99.8 | |
| 2.1% | SDBioline | 49.3 | 99.7 | |
| 5.1% | DDTD | 78.7 | 99.3 | |
| 5.1% | GADx | 80.7 | 99.6 | |
| 5.1% | SDBioline | 70.9 | 99.3 | |
| 10.1% | DDTD | 88.5 | 98.5 | |
| 10.1% | GADx | 89.7 | 99.1 | |
| 10.1% | SDBioline | 83.6 | 98.5 | |
| Prevalence | **Test** | **Expected FP per 1000** | **95% CrI** | |
| 1% | SDBioline | 19.0 | 13.8 – 25.1 | |
| 5% | SDBioline | 18.2 | 13.2 – 24.1 | |
| 10% | SDBioline | 17.2 | 12.5 – 22.9 | |
| 1% | DDTD | 12.5 | 8.2 – 17.6 | |
| 5% | DDTD | 12.0 | 7.9 – 16.9 | |
| 10% | DDTD | 11.3 | 7.5 – 16.0 | |
| 1% | GADx | 11.7 | 6.7 – 17.2 | |
| 5% | GADx | 11.2 | 6.4 – 16.5 | |
| 10% | GADx | 10.6 | 6.1 – 15.7 | |

**Table S1c. Model 3 – Decision metrics for Ov16 RDTs**

Posterior median sensitivity (Se) and specificity (Sp), probability of meeting WHO thresholds (Se ≥89% for stop-MDA decisions, Sp ≥99.8%), predictive values (PPV, NPV) at representative elimination-setting prevalences, and expected number of false positives per 1,000 tested.

| Test | Median Se (%) | P(Se ≥89%) | Median Sp (%) | P(Sp ≥99.8%) |
| --- | --- | --- | --- | --- |
| SDBioline | 92.7 | 0.955 | 98.0 | 0.000 |
| DDTD | 88.4 | 0.418 | 98.7 | 0.000 |
| GADx | 92.8 | 0.968 | 98.5 | 0.000 |
| Prevalence | **Test** | **PPV (%)** | **NPV (%)** | |
| 0.6% | DDTD | 28.4 | 99.9 | |
| 0.6% | GADx | 27.8 | 100.0 | |
| 0.6% | SDBioline | 21.7 | 100.0 | |
| 1.1% | DDTD | 42.2 | 99.9 | |
| 1.1% | GADx | 41.5 | 99.9 | |
| 1.1% | SDBioline | 33.8 | 99.9 | |
| 2.1% | DDTD | 58.5 | 99.7 | |
| 2.1% | GADx | 57.8 | 99.8 | |
| 2.1% | SDBioline | 49.6 | 99.8 | |
| 5.1% | DDTD | 77.9 | 99.4 | |
| 5.1% | GADx | 77.4 | 99.6 | |
| 5.1% | SDBioline | 71.1 | 99.6 | |
| 10.1% | DDTD | 88.1 | 98.7 | |
| 10.1% | GADx | 87.8 | 99.2 | |
| 10.1% | SDBioline | 83.8 | 99.2 | |
| Prevalence | **Test** | **Expected FP per 1000** | **95% CrI** | |
| 1% | SDBioline | 20.0 | 14.5 – 26.3 | |
| 5% | SDBioline | 19.2 | 13.9 – 25.3 | |
| 10% | SDBioline | 18.2 | 13.1 – 24.0 | |
| 1% | DDTD | 13.3 | 9.0 – 18.5 | |
| 5% | DDTD | 12.7 | 8.7 – 17.7 | |
| 10% | DDTD | 12.1 | 8.2 – 16.8 | |
| 1% | GADx | 14.4 | 9.3 – 20.3 | |
| 5% | GADx | 13.8 | 9.0 – 19.5 | |
| 10% | GADx | 13.1 | 8.5 – 18.5 | |

**Table S1d. Model 4 – Decision metrics for Ov16 RDTs**

Posterior median sensitivity (Se) and specificity (Sp), probability of meeting WHO thresholds (Se ≥89% for stop-MDA decisions, Sp ≥99.8%), predictive values (PPV, NPV) at representative elimination-setting prevalences, and expected number of false positives per 1,000 tested.

| Test | Median Se (%) | P(Se ≥89%) | Median Sp (%) | P(Sp ≥99.8%) |
| --- | --- | --- | --- | --- |
| SDBioline | 86.1 | 0.054 | 98.4 | 0.000 |
| DDTD | 87.1 | 0.263 | 97.7 | 0.000 |
| GADx | 89.9 | 0.674 | 99.0 | 0.000 |
| Prevalence | **Test** | **PPV (%)** | **NPV (%)** | |
| 0.6% | DDTD | 18.8 | 99.9 | |
| 0.6% | GADx | 34.7 | 99.9 | |
| 0.6% | SDBioline | 24.0 | 99.9 | |
| 1.1% | DDTD | 29.9 | 99.9 | |
| 1.1% | GADx | 49.4 | 99.9 | |
| 1.1% | SDBioline | 36.7 | 99.8 | |
| 2.1% | DDTD | 45.1 | 99.7 | |
| 2.1% | GADx | 65.4 | 99.8 | |
| 2.1% | SDBioline | 52.8 | 99.7 | |
| 5.1% | DDTD | 67.3 | 99.3 | |
| 5.1% | GADx | 82.5 | 99.5 | |
| 5.1% | SDBioline | 73.7 | 99.2 | |
| 10.1% | DDTD | 81.2 | 98.5 | |
| 10.1% | GADx | 90.8 | 98.9 | |
| 10.1% | SDBioline | 85.4 | 98.4 | |
| Prevalence | **Test** | **Expected FP per 1000** | **95% CrI** | |
| 1% | SDBioline | 16.3 | 11.4 – 22.3 | |
| 5% | SDBioline | 15.7 | 11.0 – 21.4 | |
| 10% | SDBioline | 14.8 | 10.4 – 20.2 | |
| 1% | DDTD | 22.5 | 16.9 – 29.0 | |
| 5% | DDTD | 21.6 | 16.2 – 27.9 | |
| 10% | DDTD | 20.4 | 15.3 – 26.4 | |
| 1% | GADx | 10.1 | 5.6 – 15.4 | |
| 5% | GADx | 9.7 | 5.4 – 14.7 | |
| 10% | GADx | 9.2 | 5.1 – 14.0 | |

**Table S1e. Model 5 – Decision metrics for Ov16 RDTs**

Posterior median sensitivity (Se) and specificity (Sp), probability of meeting WHO thresholds (Se ≥89% for stop-MDA decisions, Sp ≥99.8%), predictive values (PPV, NPV) at representative elimination-setting prevalences, and expected number of false positives per 1,000 tested.

| Test | Median Se (%) | P(Se ≥89%) | Median Sp (%) | P(Sp ≥99.8%) |
| --- | --- | --- | --- | --- |
| SDBioline | 86.9 | 0.114 | 98.4 | 0.000 |
| DDTD | 87.0 | 0.248 | 98.7 | 0.000 |
| GADx | 89.9 | 0.675 | 98.9 | 0.000 |
| Prevalence | **Test** | **PPV (%)** | **NPV (%)** | |
| 0.6% | DDTD | 29.4 | 99.9 | |
| 0.6% | GADx | 32.5 | 99.9 | |
| 0.6% | SDBioline | 24.4 | 99.9 | |
| 1.1% | DDTD | 43.4 | 99.9 | |
| 1.1% | GADx | 47.0 | 99.9 | |
| 1.1% | SDBioline | 37.3 | 99.9 | |
| 2.1% | DDTD | 59.6 | 99.7 | |
| 2.1% | GADx | 63.1 | 99.8 | |
| 2.1% | SDBioline | 53.4 | 99.7 | |
| 5.1% | DDTD | 78.7 | 99.3 | |
| 5.1% | GADx | 81.1 | 99.5 | |
| 5.1% | SDBioline | 74.2 | 99.3 | |
| 10.1% | DDTD | 88.5 | 98.5 | |
| 10.1% | GADx | 90.0 | 98.9 | |
| 10.1% | SDBioline | 85.7 | 98.5 | |
| Prevalence | **Test** | **Expected FP per 1000** | **95% CrI** | |
| 1% | SDBioline | 16.1 | 11.2 – 21.9 | |
| 5% | SDBioline | 15.4 | 10.8 – 21.0 | |
| 10% | SDBioline | 14.6 | 10.2 – 19.9 | |
| 1% | DDTD | 12.5 | 8.2 – 17.7 | |
| 5% | DDTD | 12.0 | 7.9 – 17.0 | |
| 10% | DDTD | 11.3 | 7.5 – 16.1 | |
| 1% | GADx | 11.2 | 6.5 – 16.8 | |
| 5% | GADx | 10.7 | 6.2 – 16.1 | |
| 10% | GADx | 10.1 | 5.9 – 15.2 | |

**Table S2. Site selection, sampling methods, and study population characteristics across the three country evaluations**

This table summarizes the site-selection criteria, planned and actual sampling locations, target populations, sampling approaches, and programmatic contexts of the field evaluations conducted in Ghana, Mozambique, and Benin. OEM, onchocerciasis elimination mapping; IVM, ivermectin; LF, lymphatic filariasis; IRB, institutional review board.

|  | **Ghana** | **Mozambique** | **Benin** |
| --- | --- | --- | --- |
| **Site selection criteria** | Based on the most recent available prevalence data | Based on the results of a breeding site assessment and previous Ov16 positives | The selection of districts included in this study was based on the criteria of the main survey, the serological threshold study. As such, the targeted districts have a recent seroprevalence of below or equal to 2% and mass drug administration has been implemented for more than 15 years with the possibility that transmission has been interrupted. |
| **Sites initially planned** | 6 communities in 2 districts | 2 communities in 2 districts | **Two** districts initially targeted: Tchaourou and Save |
| **Sites sampled** | 7 communities in 3 districts | 2 communities in 2 districts | A total of **3** districts were sampled during implementation |
| **Reason for change in number of sites** | Inability to reach the sample sizes. The protocol was amended and approved by the IRB for the addition of the 7th site | Not applicable | In Tchaourou, the field team faced major challenges, including empty households in targeted villages because residents had moved to Nigeria for farming during the rainy season, as well as high refusal rates.  To meet the sample target, an additional district was selected. |
| **Target population** | 1700; individuals 5 years and above; skin snip for individuals 18 years and above | The study targeted children less than 10 years (5-9 years), and adults aged 20 years or older. | Participants in this study are residents aged 5 years and above, who agreed to take part in the study on the serological threshold study. Residents are children and adults born and living in the village for at least three years. |
| **Sampling method** | Convenience sampling | At each of the two sites, 100 adults and 100 children were sampled, using convenience sampling to select them | Villages were selected by probability proportional to estimated size method, plus one additional first-line village.  In villages, a multi-stage random sample of children was enrolled with parental authorization |
| **Probability sampling procedures** | Not applicable | Not applicable | As described in the cell above |
| **Non-probability sampling procedures** | Not applicable | Not applicable | In households, where there is more than one child aged between 5 and 9 in a household, the sampling agent has the option of sampling other children in the household, provided that the sample size of 1,200 is reached in all the villages selected. |
| **Programmatic context** | Pre-stop | OEM, but previously treated with IVM for LF | Pre-Stop |
