## Supplemental Appendix 2 for "Assessing diagnostic accuracy of onchocerciasis rapid diagnostic tests using Bayesian latent class models"

**S2 Appendix. - Latent class Bayesian models**

### 1. Notation and model formulation

Let *i* = 1, …, *N* index the participants with complete results on all three Ov16 tests, and let *c*(*i*) ∈ {Benin, Ghana, Mozambique} denote the country of participant *i*. For each participant we observe the results of three index tests, *j* = 1 (SD Bioline), *j* = 2 (DDTD), and *j* = 3 (GADx):

T_ij_ ∈ {0, 1}, *j* = 1, 2, 3,

where T_ij_ = 1 denotes a positive result of test *j* for participant *i*. The true, unobserved Ov16 exposure status of participant *i* is denoted by the latent variable

D_i_ ∈ {0, 1},

and the country-specific prevalence of exposure is

π_c_ = Pr(D_i_ = 1 ∣ *c*(*i*) = *c*), *c* ∈ {Benin, Ghana, Mozambique}.

The sensitivity and specificity of test *j* are

Se_j_ = Pr(T_ij_ = 1 ∣ D_i_ = 1), Sp_j_ = Pr(T_ij_ = 0 ∣ D_i_ = 0).

Consistent with the main text, Se_j_ and Sp_j_ are assumed common across countries (properties of the assay itself), while π_c_ is allowed to vary by country (a property of the population sampled).

Under the standard conditional independence assumption for latent class models (20–22), the three test results are independent of one another given the true latent status D_i_:

Pr(T_i1_ = t_1_, T_i2_ = t_2_, T_i3_ = t_3_ ∣ D_i_) = ∏^3^_j = 1_ Pr(T_ij_ = t_j_ ∣ D_i_).

Marginalising over the unobserved status D_i_, the (marginal) probability of observing pattern (t_1_, t_2_, t_3_) for a participant in country *c* is

Pr(T_i1_=t_1_,T_i2_=t_2_,T_i3_=t_3_) = π_c_ ∏^3^_j_ Se_j_^tⱼ^(1−Se_j_)^1−tⱼ^

  + (1−π_c_) ∏^3^_j_ (1−Sp_j_)^tⱼ^ Sp_j_^1−tⱼ^.

Because there are three binary tests, there are 2^3^ = 8 possible response patterns per participant. Let ℰ = {0,1}^3^ denote the set of these patterns *r* = (r_1_, r_2_, r_3_), and for country *c* let n_c,r_ denote the observed number of participants exhibiting pattern *r*, with N_c_ = Σ_r∈ℰ_ n_c,r_. The observed cell counts within each country follow a multinomial distribution:

(n_c,r_)_r∈ℰ_ ~ Multinomial(N_c_ ; (p_c,r_)_r∈ℰ_),

where p_c,r_ is the expression above evaluated at (t_1_, t_2_, t_3_) = *r*. The full likelihood, pooling all three countries, is

L( π_Benin_, π_Ghana_, π_Moz_, {Se_j_, Sp_j_}_j=1,2,3_ ∣ data ) = ∏_c_ ∏_r∈ℰ_ p_c,r_^n^c,r.

This is the standard Hui–Walter–type formulation for a single population (here, extended to three countries with a shared set of test parameters), on which Models 1–5 in the main text are all based; the models differ only in the prior distributions assigned to the parameters, described in Section 3.

### 2. Prior distributions

Table A summarises the prior distributions assigned to each parameter in Models 1–3 (the models reported in Table 1 of the main text). Models 4 and 5 use the same priors as Model 2, differing only in how a positive DDTD result is defined (Table 2 of the main text). Noninformative Uniform(0,1) priors were assigned to the sensitivity and specificity of both novel RDTs (DDTD, GADx) in every model, so that their evaluation would not be influenced by external assumptions. An informative prior for at least one test (SD Bioline) helps ensure identifiability of the two latent classes and to prevent label-switching during MCMC sampling (20).

| **Parameter** | **Model 1 (manufacturer prior)** | **Model 2 (field prior); also Models 4–5** | **Model 3 (no prior)** |
| --- | --- | --- | --- |
| π (Benin, Ghana, Mozambique) | Uniform(0,1), each country | Uniform(0,1), each country | Uniform(0,1), each country |
| Se, SD Bioline | Beta(53.540, 12.477) mean = 81.1% | Beta(74.718, 25.172) mean = 74.8% | Uniform(0,1) |
| Sp, SD Bioline | Beta(217.275, 2.195) mean = 99.0% | Beta(119.103, 1.691) mean = 98.6% | Uniform(0,1) |
| Se, Sp — DDTD | Uniform(0,1) | Uniform(0,1) | Uniform(0,1) |
| Se, Sp — GADx | Uniform(0,1) | Uniform(0,1) | Uniform(0,1) |

*Table A. Prior distributions by model. π, prevalence; Se, sensitivity; Sp, specificity.*

Which also considers **Models 4** and **Model 5** which retain the same prior distributions as Model 2 (Table A) and differ only in how a positive DDTD result is defined. Model 2 uses the manufacturer-specified definition, requiring both the T1 and T2 lines to be positive. Model 4 redefines a positive DDTD result as either line, T1 or T2, being positive. Model 5 redefines a positive DDTD result as the T1 line alone being positive, regardless of the T2 result. In each case, only the classification rule applied to the DDTD test result is changed; the likelihood, prevalence structure, and priors for SD Bioline and GADx are unchanged from Model 2.

The manufacturer prior mean sensitivity (81.1%) and specificity (99.0%) were taken from Abbott's reported performance of the SD Bioline Ov16 RDT (15, 16). The field prior mean sensitivity (74.8%) and specificity (98.6%) were taken from field-based estimates reported by Hotterbeekx et al. (5).

For each of the four quantities, the corresponding mean was combined with the reported 95% confidence interval to solve for the shape parameters (*a*, *b*) of a Beta(*a*, *b*) distribution using the *findbeta()* function of the *PriorGen* R package (27), which implements the elicitation method of Branscum, Gardner & Johnson (28). The resulting shape parameters are given in Table A; their implied means reproduce the manufacturer- and field-based point estimates exactly, and the corresponding 2.5th/97.5th percentiles of each Beta distribution reproduce the reported 95% confidence intervals.

Posterior distributions were obtained by Markov Chain Monte Carlo (MCMC) simulation with 10,000 iterations per chain; the first 4,000 iterations were discarded as burn-in and the remaining 6,000 iterations were retained to construct posterior summaries (medians and 95% credible intervals). Convergence was assessed using trace plots and the Gelman–Rubin statistic across multiple chains, with *R̂* < 1.1 taken as evidence of adequate convergence for all model parameters.

### 3. Modelling assumptions

**Conditional independence**

Test results are assumed independent of one another given the true latent exposure status (Section 2). This is plausible here because the three assays differ in antigen target, platform, and specimen pathway: the two novel RDTs were performed on whole blood at the point of care, while SD Bioline was performed on dried blood spots eluted separately in the laboratory, limiting shared operational sources of correlated error. A residual source of dependence cannot be excluded, since all three tests ultimately detect the same underlying host IgG4 antibody response. If present, such dependence is expected to be positive, and positive conditional dependence is known to cause latent class models to overstate both sensitivity and specificity (20); reported estimates should therefore be interpreted as upper bounds on true test performance.

**Pooling of test accuracy across countries**

Sensitivity and specificity are treated as properties of each assay, conditional on latent exposure status, and are assumed constant across the three study countries; only prevalence is allowed to vary by country. This is supported by the use of identical test lots and protocols across sites. Sample size, and in particular the very low prevalence observed in Benin (Table 1), would not support estimation of country-specific accuracy parameters within this design.

Only participants with valid results on all three tests contribute to the multinomial cell counts n_c,r_ used in the likelihood (Section 2); participants with one or more missing or invalid test results were excluded from this analysis.

**Identifiability**

A three-test, two-class model of this kind is only weakly identified from the data alone. An informative prior on at least one test (here, SD Bioline, for which external manufacturer and field evidence exists) helps anchor the labelling of the two latent classes and stabilise estimation; the two novel RDTs were left with noninformative Uniform(0,1) priors throughout.

### 4. Convergence of the MCMC algorithms

Trace plots for all model parameters across Models 1–5 are provided as supplementary material in a compressed archive (S_trace_plots.zip), containing sensitivity and specificity trace plots for each of the three tests under each model specification (e.g. m1-SD-sen.tiff, m1-SD-spec.tiff, m2-DDTD-sen.tiff, and so on through Model 5). Across all models, chains show good mixing and convergence, consistent with the Gelman–Rubin diagnostics reported above. This is particularly evident for Model 3, which used noninformative Uniform(0,1) priors on all sensitivity and specificity parameters, where trace plots show no evidence of label switching between the latent classes.
