## Supplementary figures and images for "Assessing diagnostic accuracy of onchocerciasis rapid diagnostic tests using Bayesian latent class models"

### m1-DDTD-sen.tiff

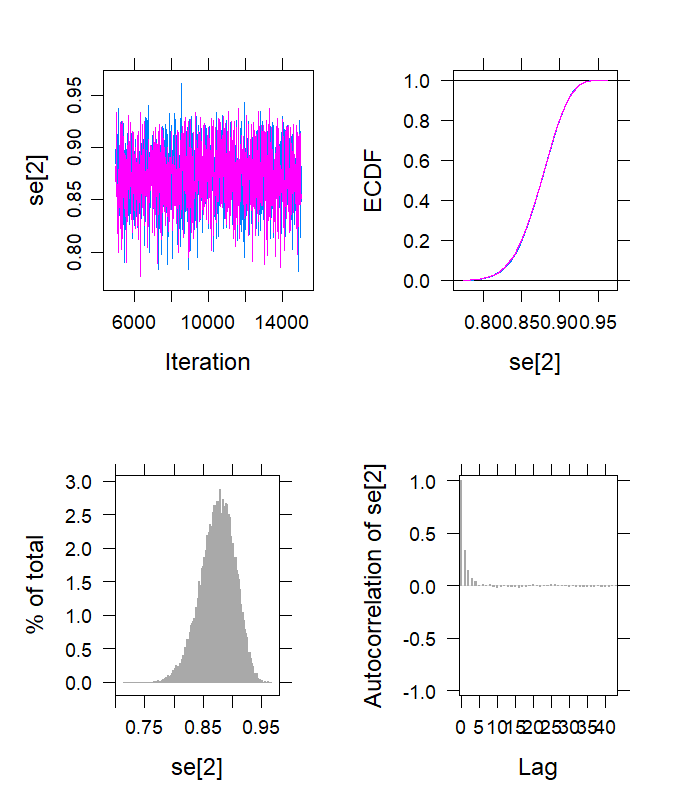

### m1-DDTD-spec.tiff

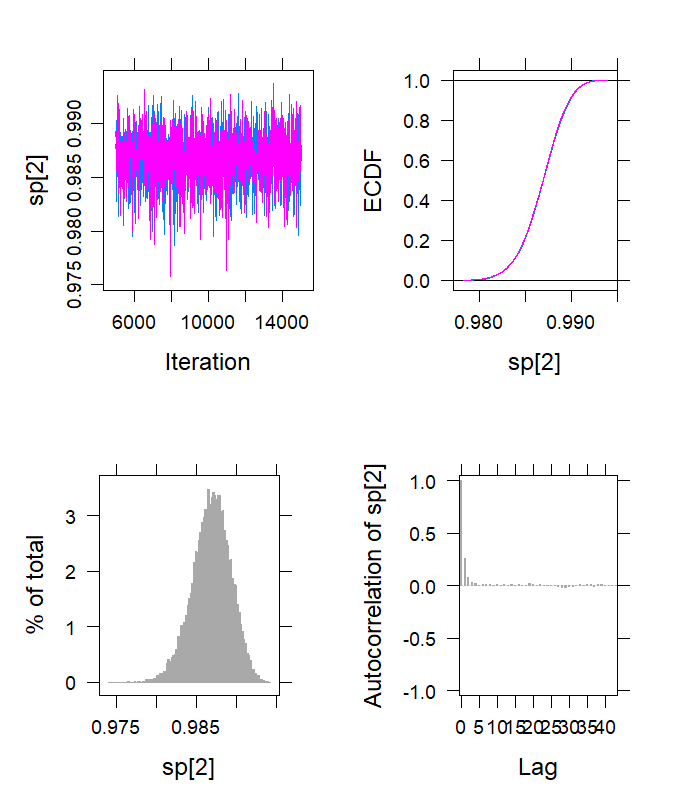

### m1-GADx-sen.tiff

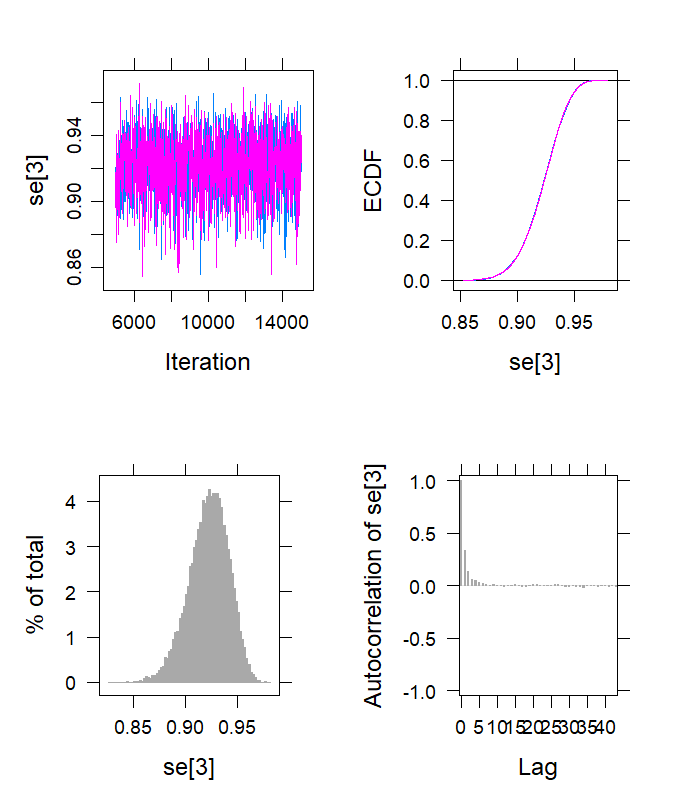

### m1-GADx-spec.tiff

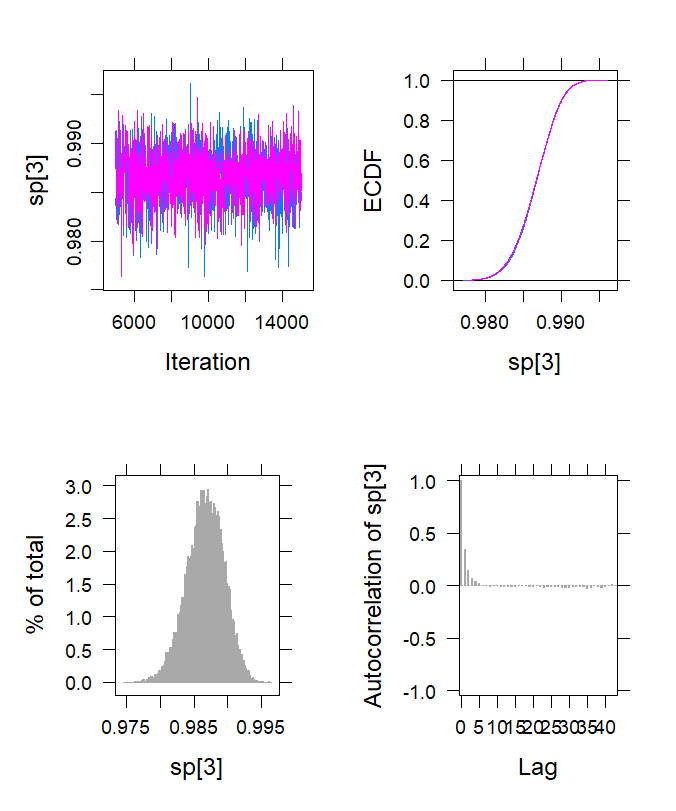

### m1-SD-sen.tiff

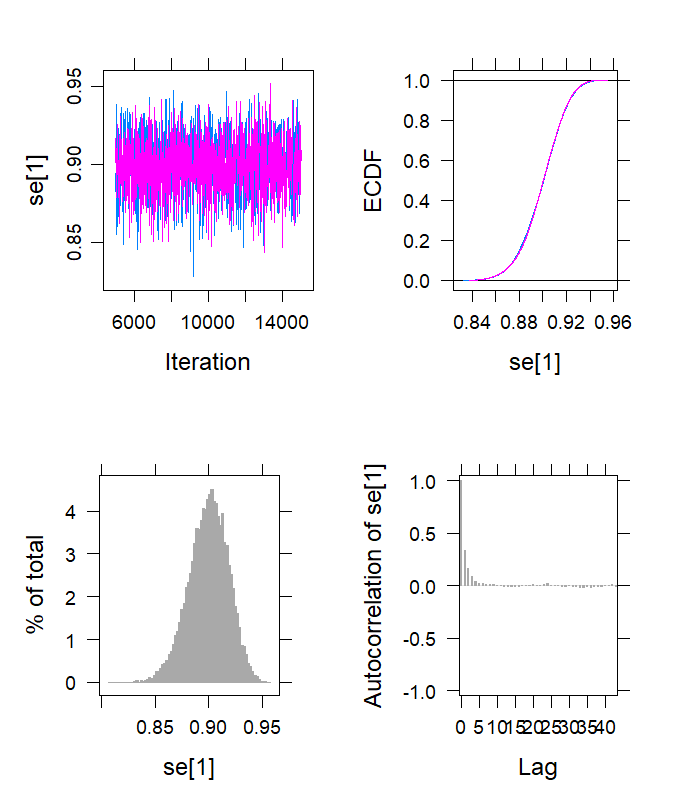

### m1-SD-spec.tiff

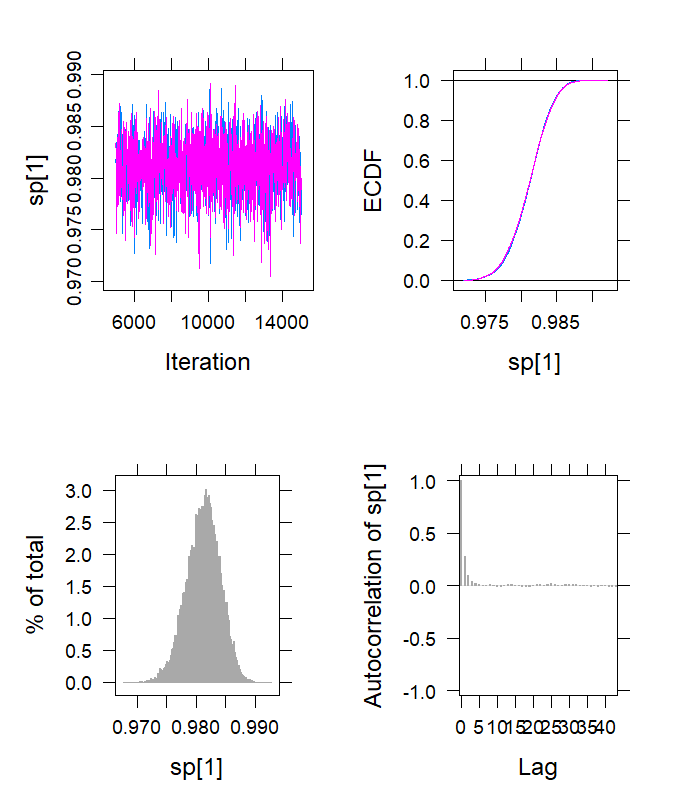

### m2-DDTD-sen.tiff

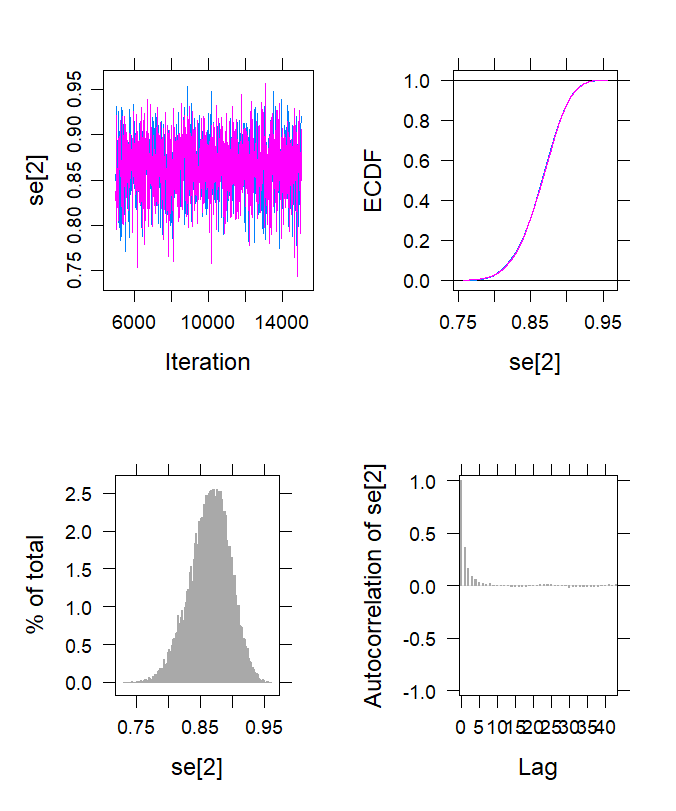

### m2-DDTD-spec.tiff

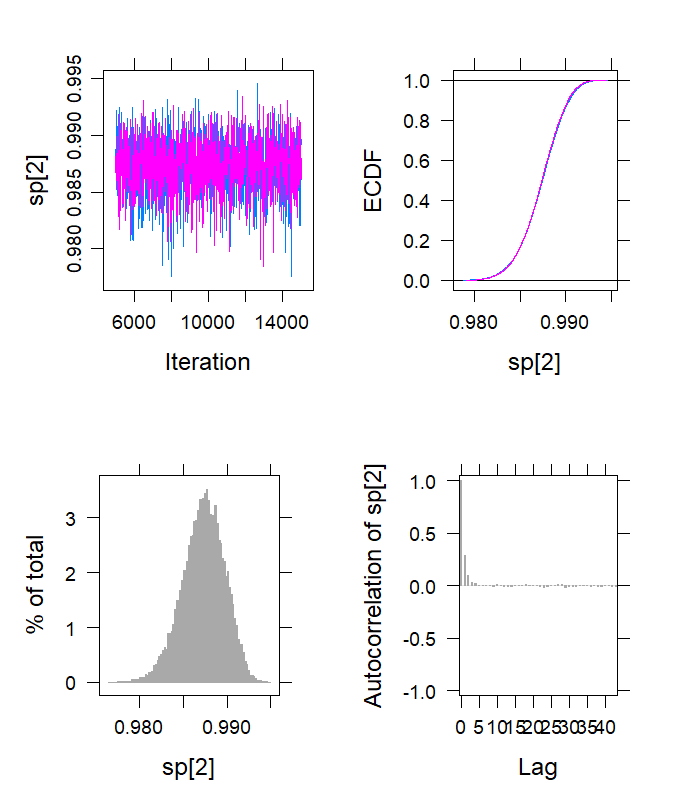

### m2-GADx-sen.tiff

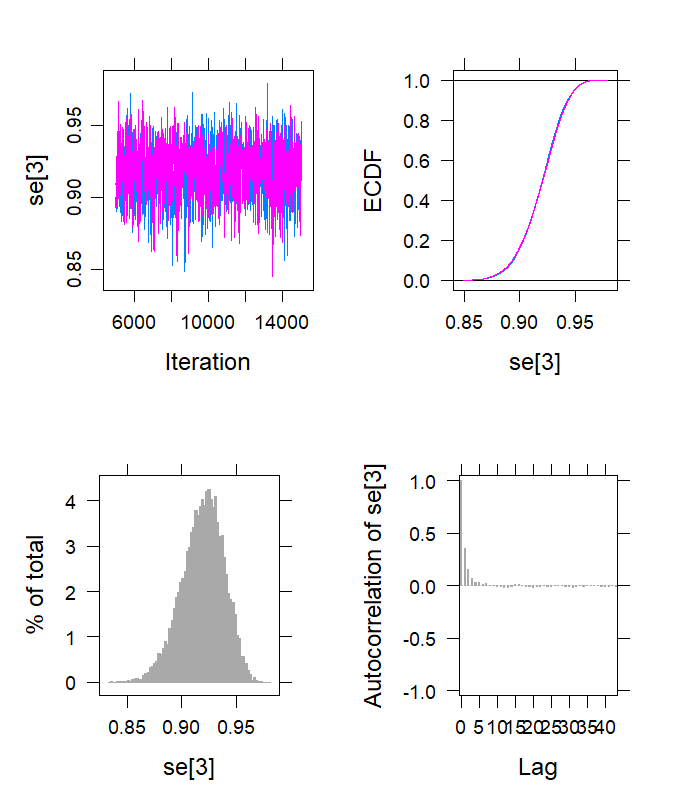

### m2-GADx-spec.tiff

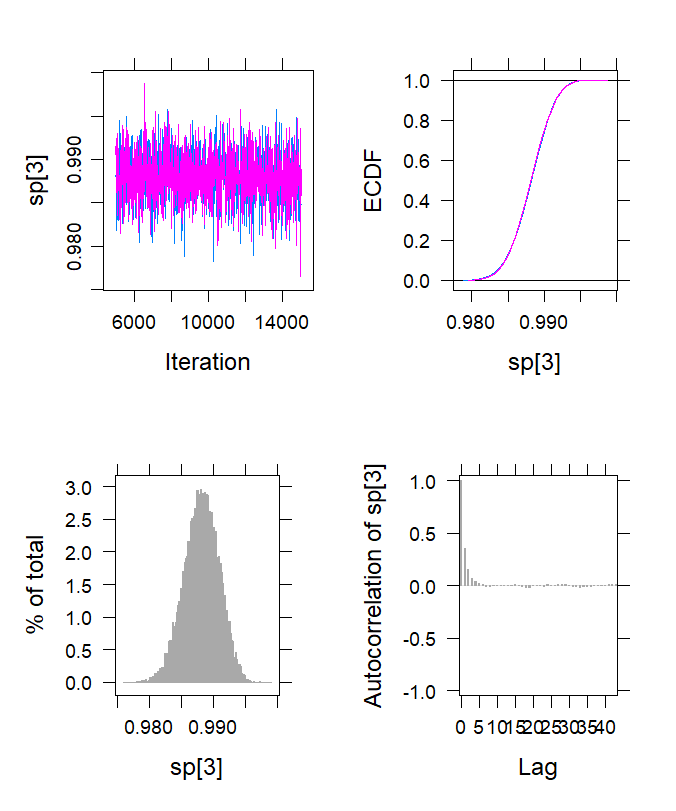

### m2-SD-sen.tiff

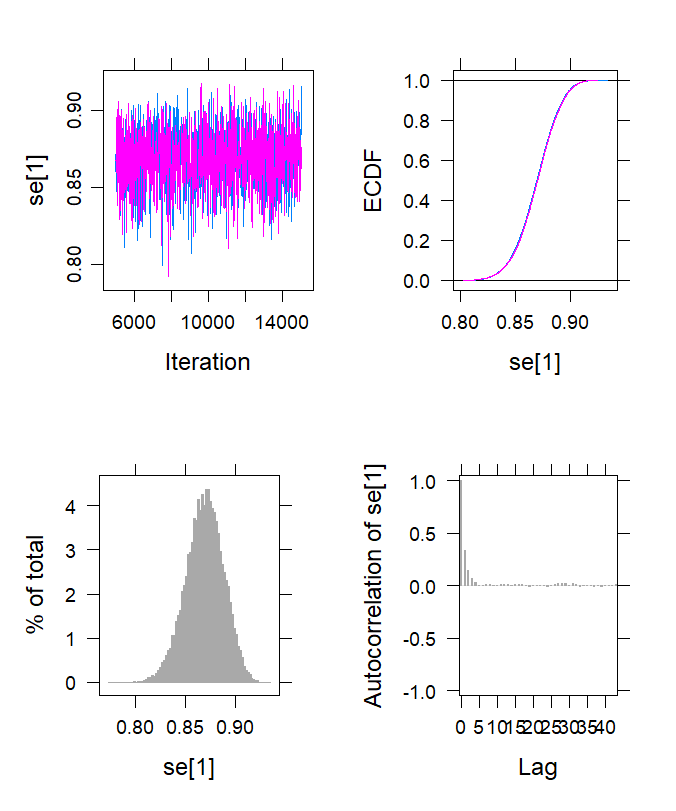

### m2-SD-spec.tiff

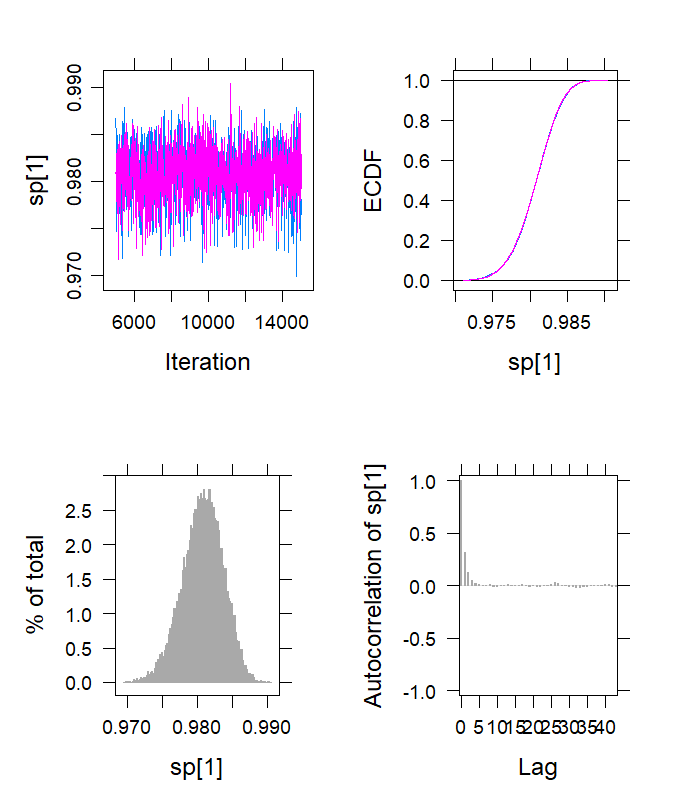

### m3-DDTD-sen.tiff

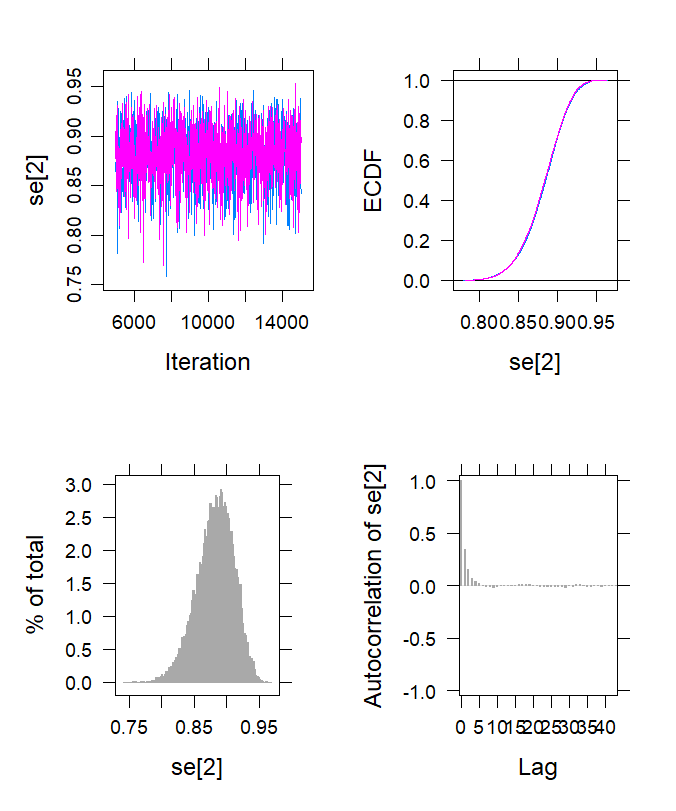

### m3-DDTD-spec.tiff

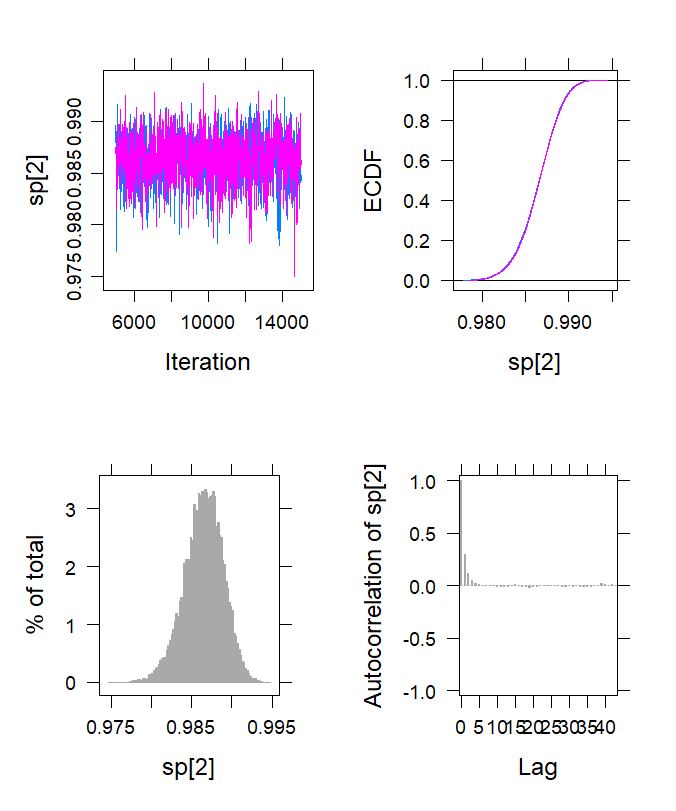

### m3-GADx-sen.tiff

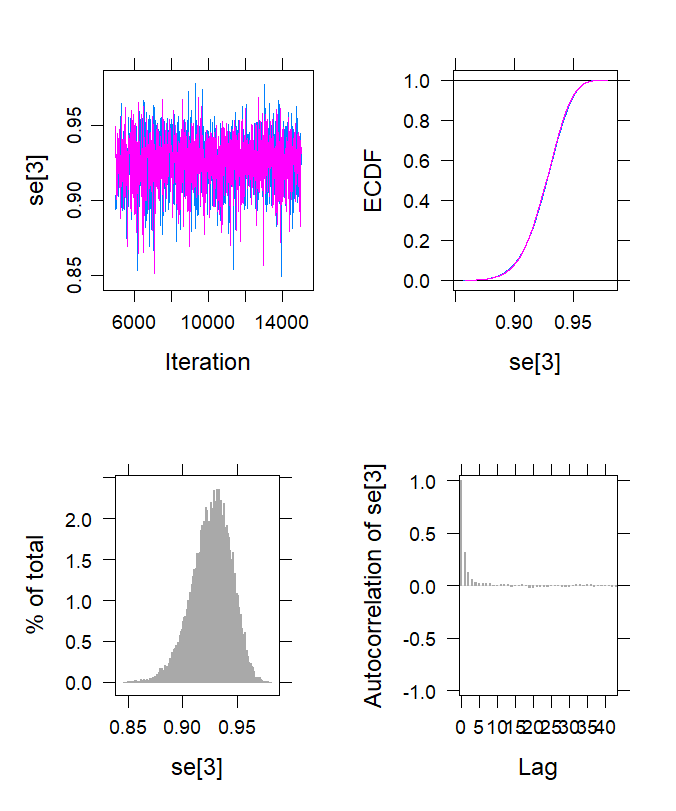

### m3-GADx-spec.tiff

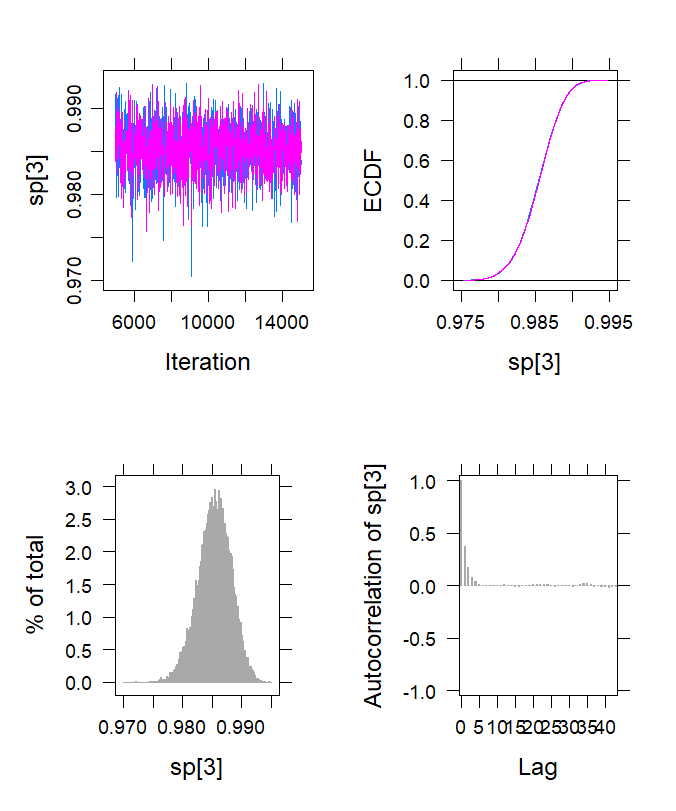

### m3-SD-sen.tiff

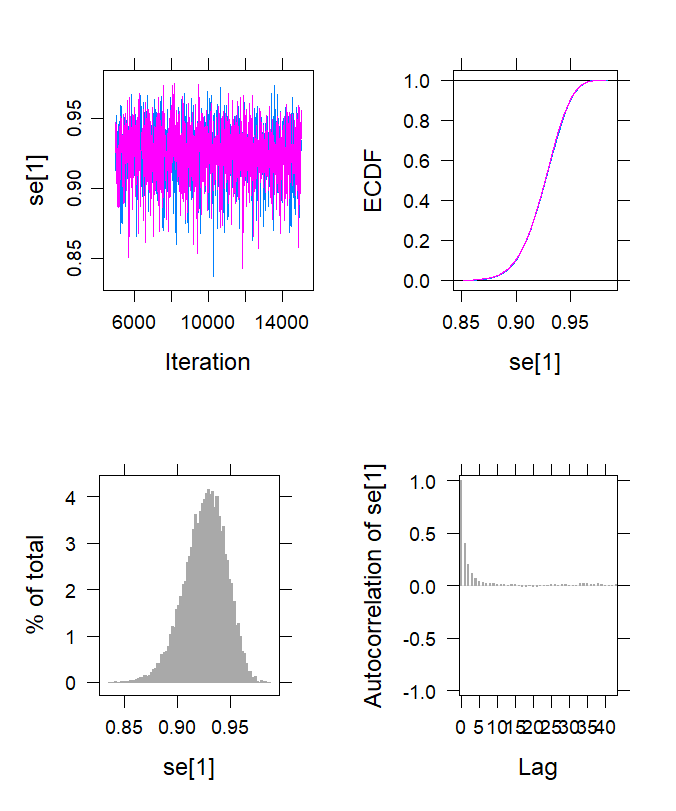

### m3-SD-spec.tiff

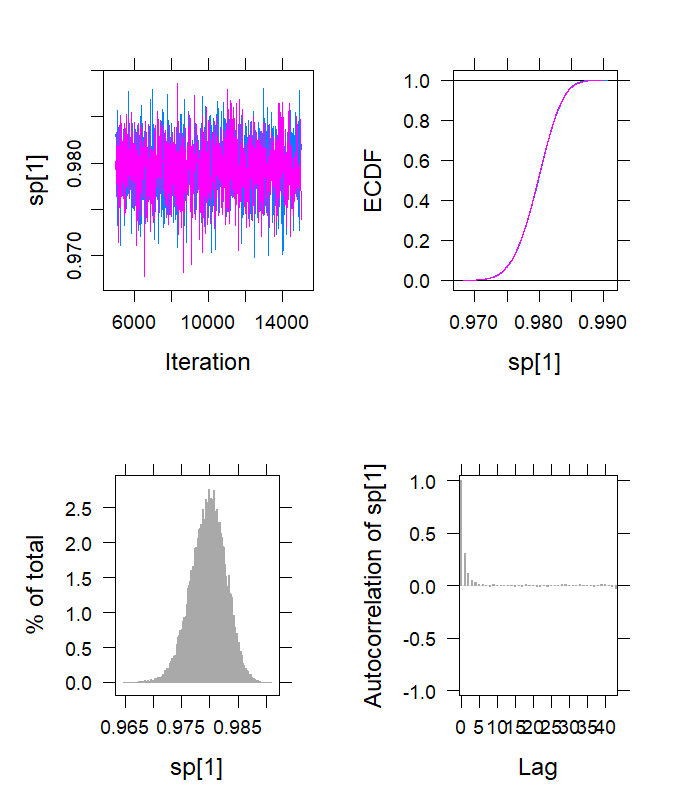

### m4-DDTD-sen.tiff

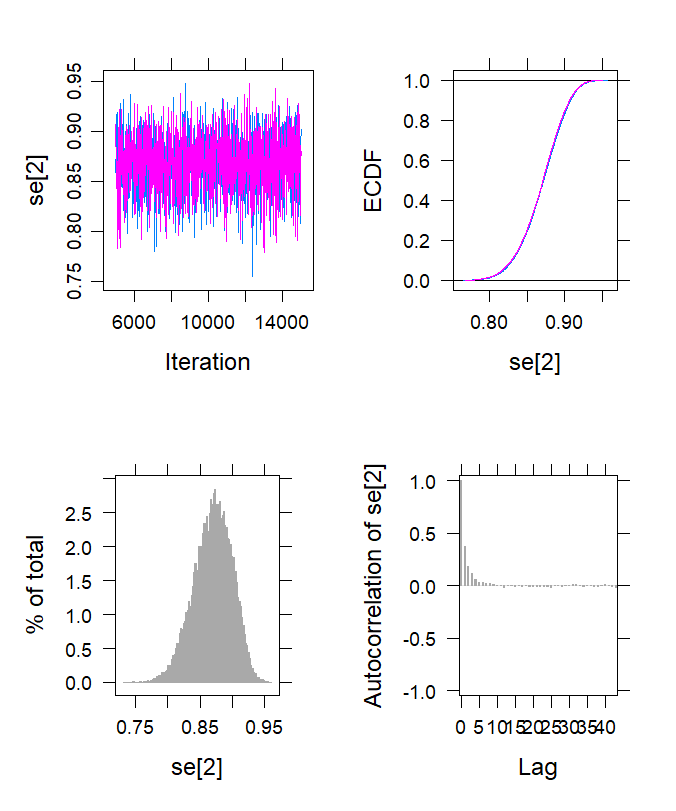

### m4-DDTD-spec.tiff

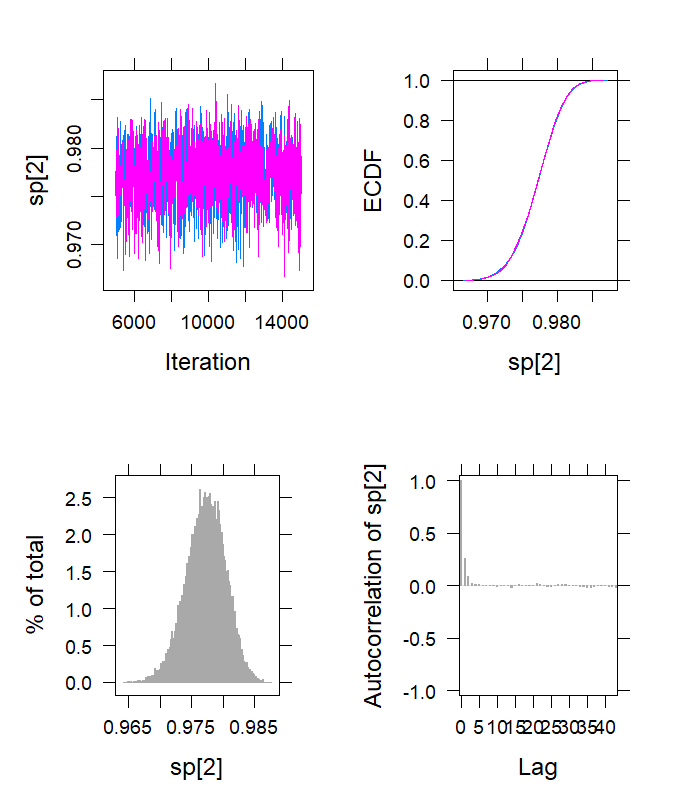

### m4-GADx-sen.tiff

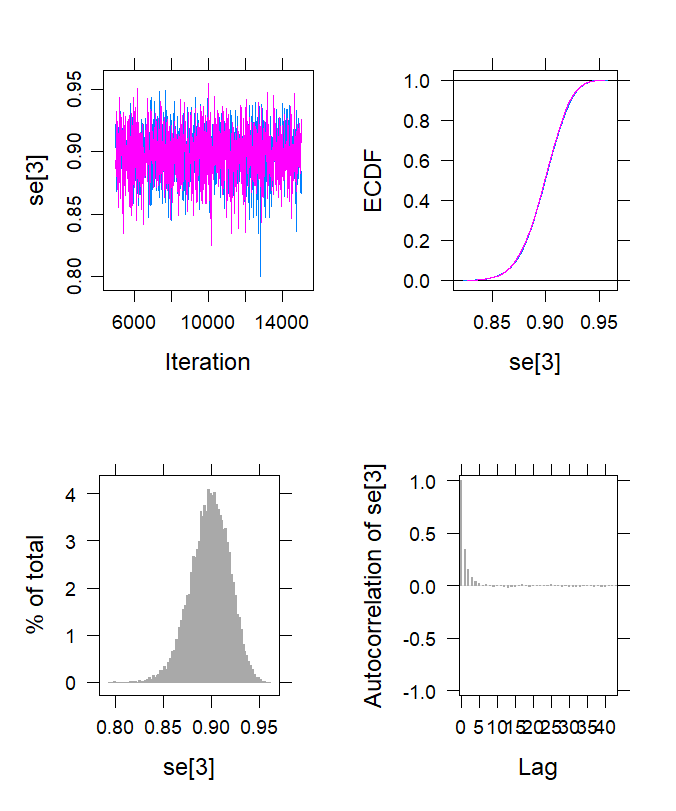

### m4-GADx-spec.tiff

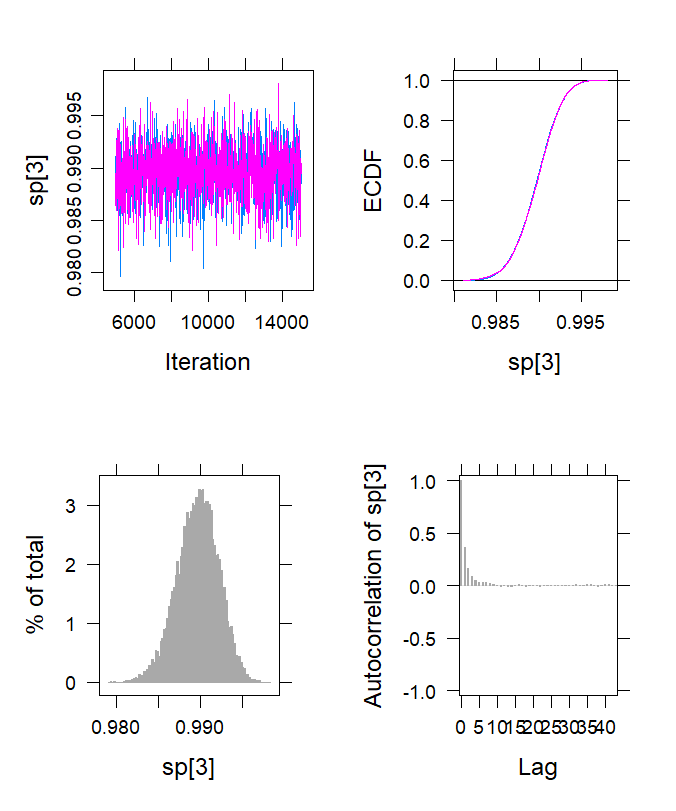

### m4-SD-sen.tiff

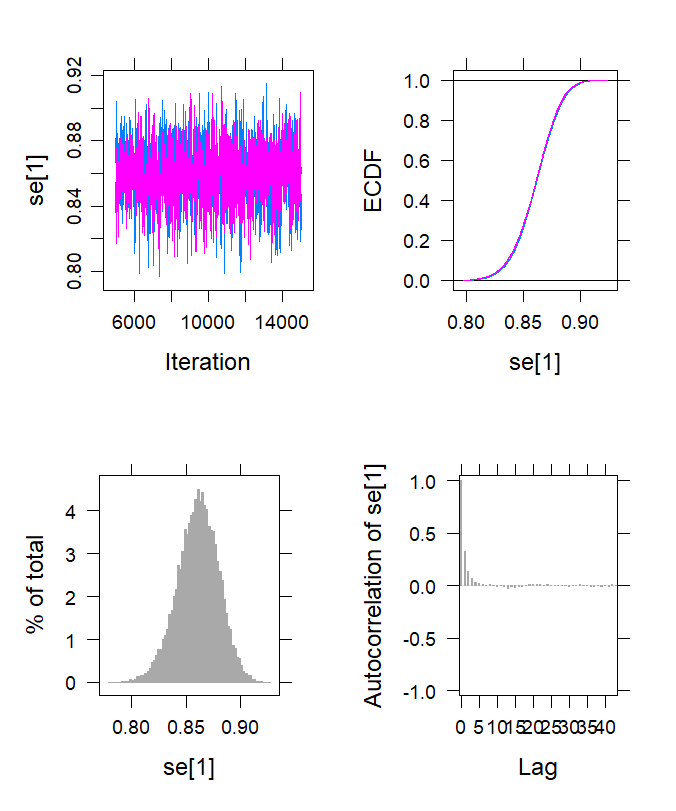

### m4-SD-spec.tiff

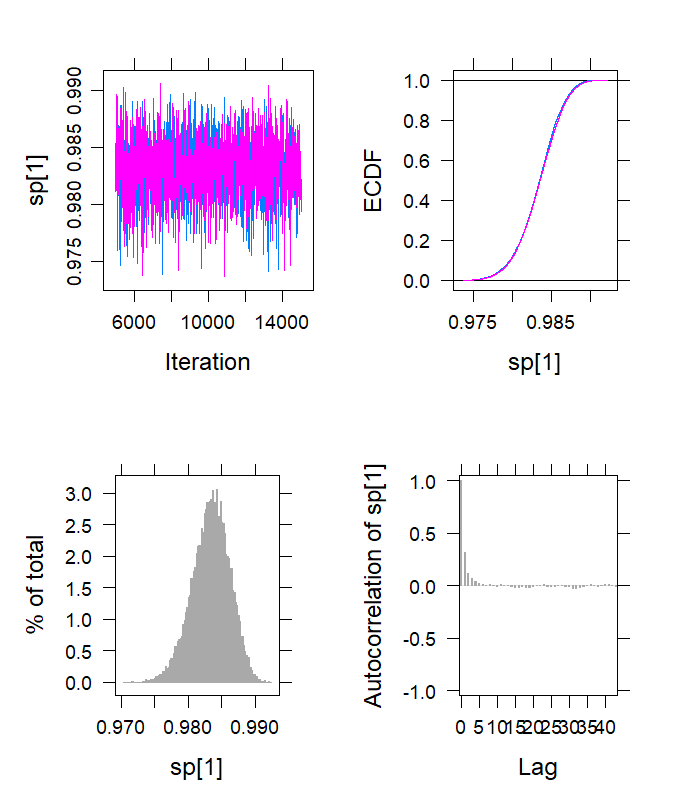

### m5-DDTD-sen.tiff

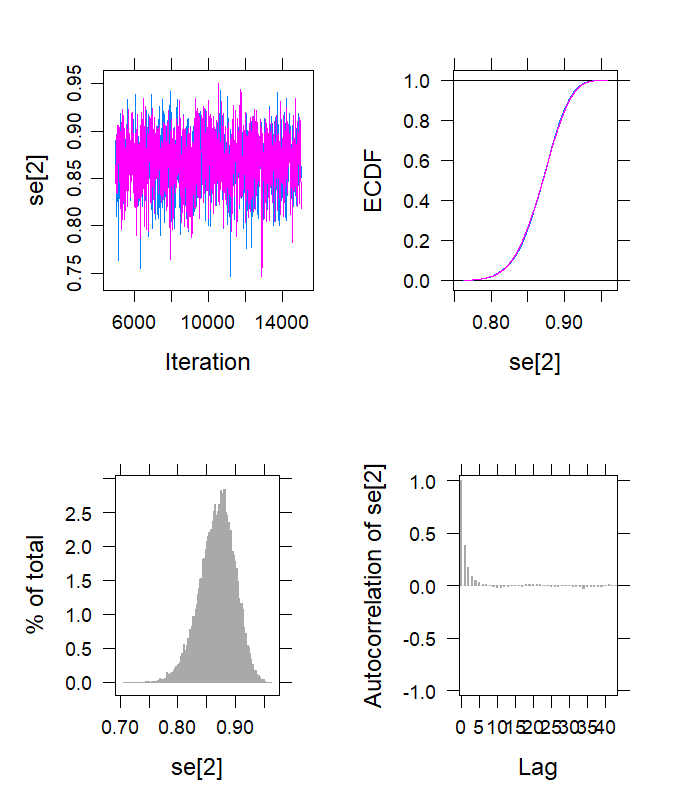

### m5-DDTD-spec.tiff

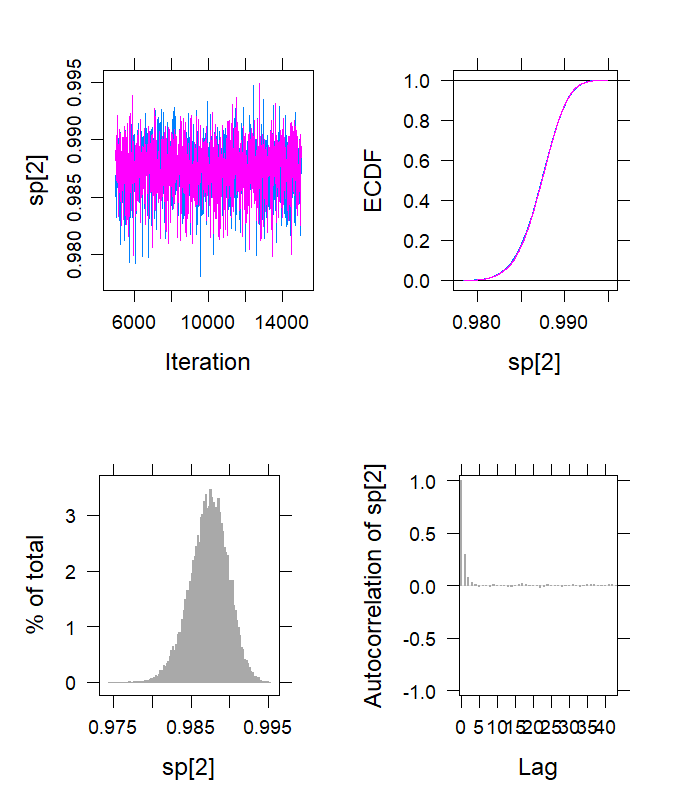

### m5-GADx-sen.tiff

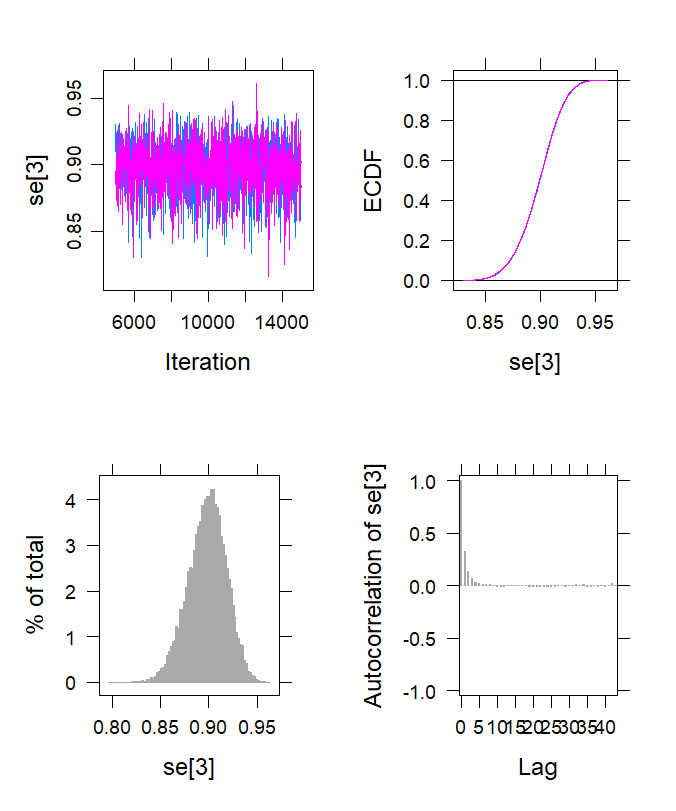

### m5-GADx-spec.tiff

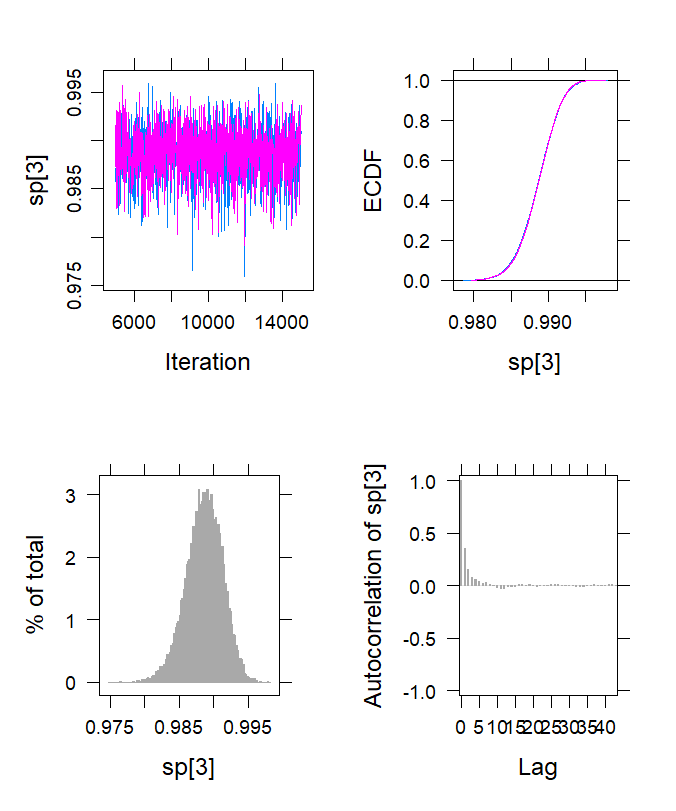

### m5-SD-sen.tiff

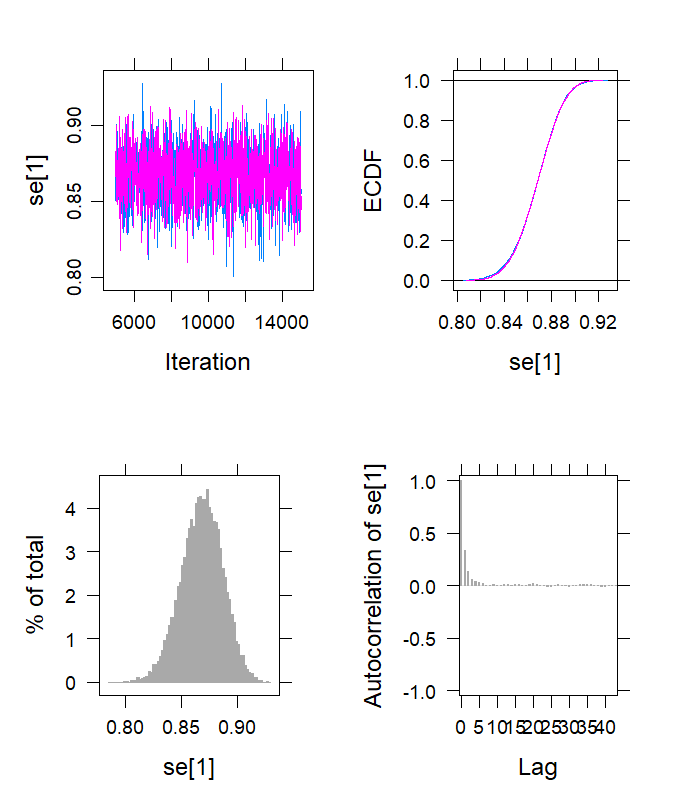

### m5-SD-spec.tiff

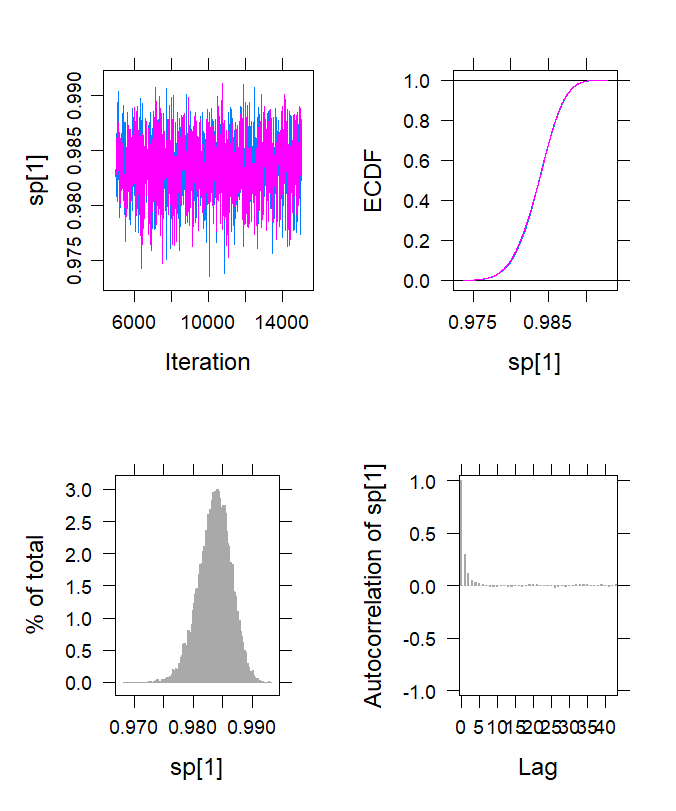
